# The bidirectional effects between cognitive ability and brain morphology: A life course Mendelian randomization analysis

**DOI:** 10.1101/2023.11.17.23297145

**Authors:** Roxanna Korologou-Linden, Isabel K. Schuurmans, Charlotte A.M. Cecil, Tonya White, Tobias Banaschewski, Arun L.W. Bokde, Sylvane Desrivières, Antoine Grigis, Hugh Garavan, Penny Gowland, Andreas Heinz, Rüdiger Brühl, Jean-Luc Martinot, Marie-Laure Paillère Martinot, Eric Artiges, Frauke Nees, Dimitri Papadopoulos Orfanos, Tomáš Paus, Luise Poustka, Nathalie Holz, Juliane H. Fröhner, M Smolka, Henrik Walter, Jeanne Winterer, Robert Whelan, Gunter Schumann, Laura D Howe, Yoav Ben-Shlomo, Neil M Davies, Emma L Anderson

**Author notes:** Joint last authors with equal contributions (Neil M Davies, Emma L Anderson).

## Abstract

**Introduction:** Little is understood about the dynamic interplay between brain morphology and cognitive ability across the life course. Additionally, most existing research has focused on global morphology measures such as estimated total intracranial volume, mean thickness, and total surface area.

**Methods:** Mendelian randomization was used to estimate the bidirectional effects between cognitive ability, global and regional measures of cortical thickness and surface area, estimated total intracranial volume, total white matter, and the volume of subcortical structures (N=37,864). Analyses were stratified for developmental periods (childhood, early adulthood, mid-to-late adulthood; age range: 8-81 years).

**Results:** The earliest effects were observed in childhood and early adulthood in the frontoparietal lobes. A bidirectional relationship was identified between higher cognitive ability, larger estimated total intracranial volume (childhood, mid-to-late adulthood) and total surface area (all life stages). A thicker posterior cingulate cortex and a larger surface area in the caudal middle frontal cortex and temporal pole were associated with greater cognitive ability. Contrary, a thicker temporal pole was associated with lower cognitive ability.

**Discussion:** Stable effects of cognitive ability on brain morphology across the life course suggests that childhood is potentially an important window for intervention.

## Introduction

Individuals with higher cognitive ability are more likely to have favourable health outcomes that impact health outcomes and quality of life. Individual differences in cognitive ability have underpinnings in both (epi-)genetic and environmental exposures ^1–3^.

Family and twin studies suggest that the heritability of cognitive ability ranges from 50 to 80% ^5–7^, with several studies suggesting estimates to increase from childhood to adulthood ^8, 9^. The latest genome-wide association study (GWAS) for cognitive ability, as measured mainly through verbal-numeric tests, identified 187 single nucleotide polymorphisms (SNPs) in 248,482 participants, indicating that cognitive ability is highly polygenic ^10^. The identified variants cluster in genes expressed in synapses and genes involved in the development of the nervous system ^10^.

Findings from 37 neuroimaging studies using functional and structural data supported the hypothesis that individual differences in human cognitive ability are predicted by structural and functional differences in the parieto-frontal network (P-FIT model) ^11^. The P-FIT model associates the network of frontal, superior temporal, middle temporal, temporal, and sensory areas in the parietal and lateral occipital regions to differences in cognitive ability. This model, which aims to reflect the neurological underpinnings of cognitive ability, was updated to consider the posterior cingulate cortex and subcortical structures such as the caudate ^12^.

The association between cognitive ability and brain morphology has previously been suggested to be age-dependent by some studies, as differences in cognitive ability are associated with size differences of different brain regions across the lifespan in observational studies. In children, there is evidence that the surface area of the prefrontal and anterior cingulate cortices is associated with cognitive ability ^13–15^. However, the orbitofrontal and middle frontal brain regions are most strongly associated with cognitive ability in adolescents (ages 12-21 years) ^16^. Cognitive ability has a strong genetic and phenotypic correlation with total cortical surface area in childhood and early adulthood. Still, there is little evidence of correlation with average cortical thickness, which may be due to the distinct genetic and phenotypic origins of these two endophenotypes ^17, 18^.

It is likely that the interplay between cognitive ability and brain morphology is dynamic and may vary across the life course. Results from observational studies may merely reflect correlation and causal inference methods such as Mendelian randomization can aid in disentangling the causality or directionality of observed associations. Establishing causality is important in finding a suitable intervention and causal inference methods integrated with age-stratification analyses can aid in identifying a critical window for these interventions. Bidirectional two-sample Mendelian randomization was used to investigate 1) whether cognitive ability has a causal effect on regional and global brain structures, 2) whether any causal effects of cognitive ability on brain morphology are time-varying, using five cohorts from different stages of the life course and 3) whether brain morphology has a causal effect on cognitive ability.

## Results

The effects of SNPs associated with cognitive ability at p<5×10^-8^ on cortical thickness, surface area, the volume of subcortical structures and total white matter were estimated in five independent cohorts across the life course ^19–25^, using two-sample Mendelian randomization ^26, 27^. Univariable Mendelian randomization using a random-effects inverse variance weighted (IVW) regression was employed to estimate the causal effect of cognitive ability on structural brain measures. The causal effect estimates of cognitive ability on brain morphology were interpreted as a one standard deviation (SD) change in brain morphology per one SD increase in cognitive ability. Descriptive statistics are in Table 1.

**Table 1.** Descriptive statistics for included samples across the life course.

**Table 2.**
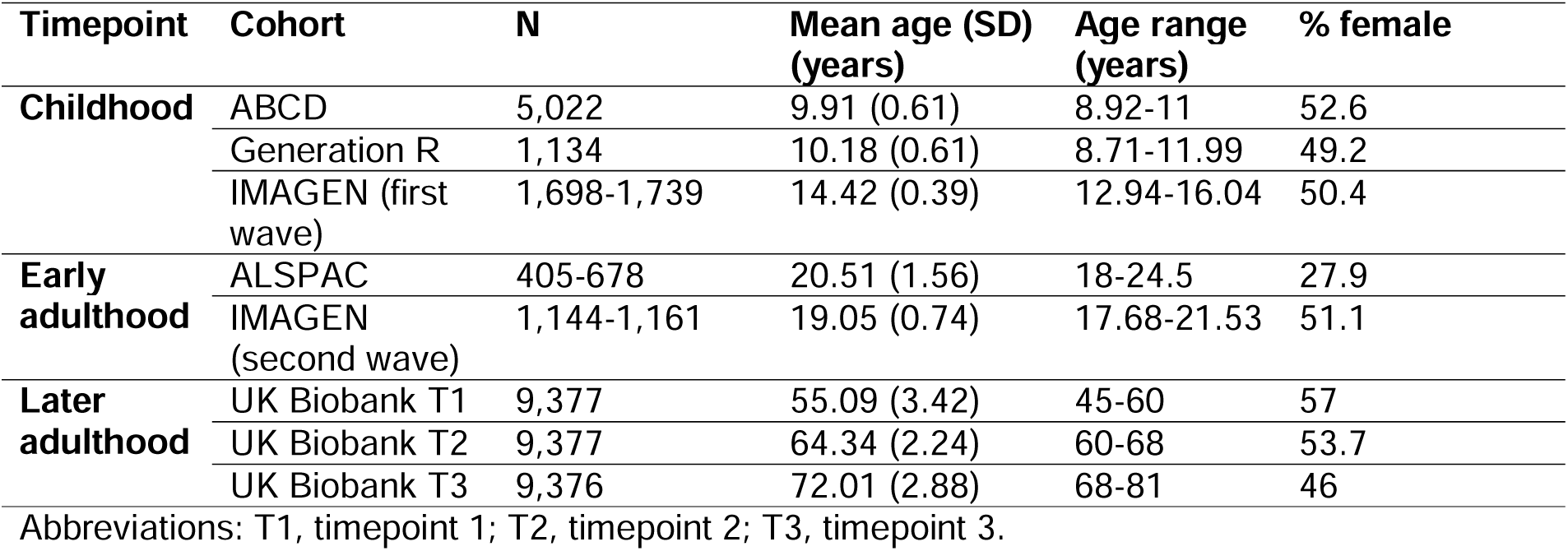
Descriptive statistics for included samples across the life course.

Forest plots for the age-stratified analyses can be found in Figures 1-3, where unadjusted p-values for the age trend have been included for each region/structure. A p-value corrected for the false discovery rate (FDR) in the age-stratified analyses is provided in Supplementary tables 4, 8, and 9. After this adjustment, there was evidence of association between cognitive ability and 13 outcomes (e.g., total surface area, estimated total intracranial volume, accumbens, superior temporal thickness, entorhinal area and superior parietal thickness). Results for the effects of cognitive ability on regional cortical thickness and surface area at each stage in the life course were visualised with a brain atlas (Figure 4), using the ggseg tool ^28^.

**Figure 1a.**
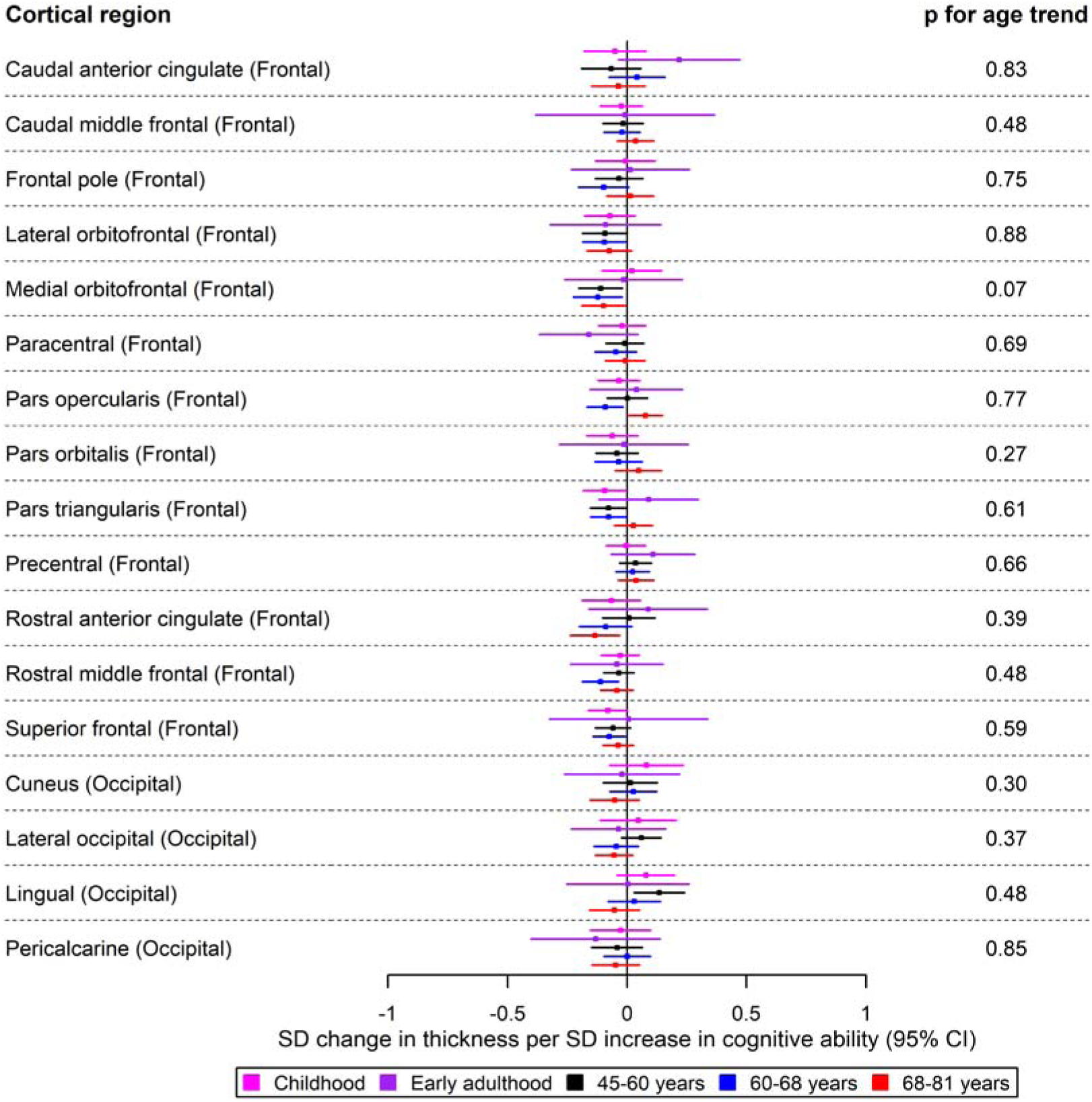
The causal effects of genetically predicted cognitive ability on the thickness of the frontal and occipital cortices at different ages across the life course (see Figure 1b for structures in the + parietal and temporal cortices, as well as mean thickness). The childhood cohorts include meta-analysed effects of three peri-pubertal cohorts: ABCD, GEN R and IMAGEN. The early adulthood cohort includes meta-analysed effects of ALSPAC and IMAGEN (second wave for data collection), and the later adulthood cohort includes UK Biobank. Effect estimates represent SD changes in thickness. Regional measures were adjusted for mean thickness. Where an effect estimate is missing, that structural measure was unavailable in that cohort.

**Figure 1b.**
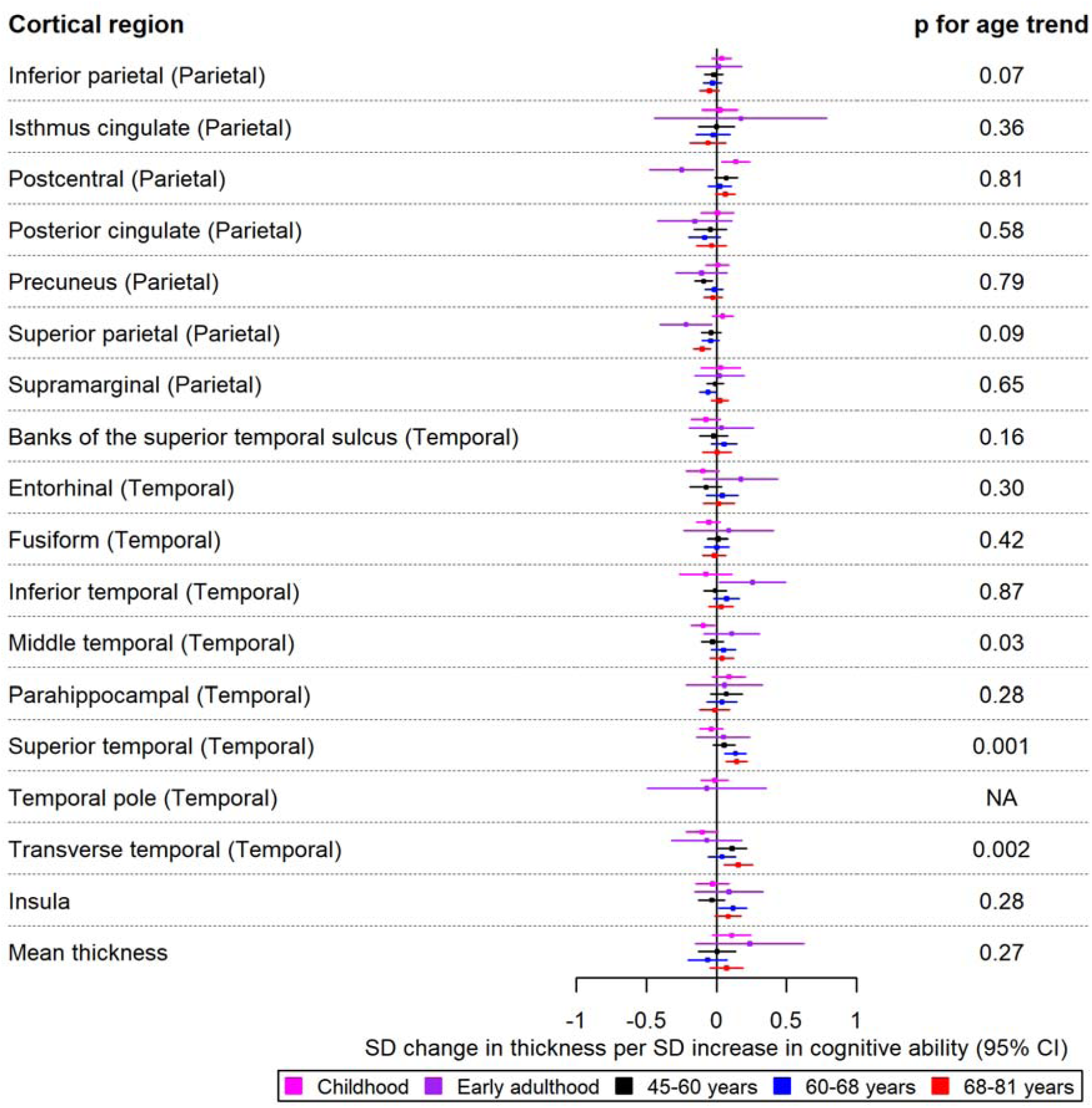
The causal effects of genetically predicted cognitive ability on the thickness of the parietal and temporal cortices at different ages across the life course. The early adulthood cohort includes meta-analysed effects of ALSPAC and IMAGEN (second wave for data collection), and the later adulthood cohort includes UK Biobank. Effect estimates represent SD changes in surface area. Regional measures were adjusted for total surface area. Where an effect estimate is missing, that structural measure was unavailable in that cohort.

Additionally, we aimed to replicate the age-stratified results in in the Enhancing Neuroimaging Genetics through Meta-analysis (ENIGMA) consortium, which is larger and more well-powered than individual cohort studies with genetic and neuroimaging data.

In the reverse direction, the causal effects of cortical thickness, surface area, and volumes of subcortical structures on cognitive ability were estimated using summary-level data from ENIGMA ^29–32^ (Figure 5). Again, univariable Mendelian randomization was employed to examine the causal effects of each brain structure on cognitive ability. All effect estimates represent an SD change in cognitive ability per standard deviation increase in brain structure. Steiger directionality tests were performed to examine whether the genetic instruments are valid for the direction of analysis that they were used in.

In the online repository^1^, detailed results with false discovery rate adjusted p-values accounting for the number of tests in the age-stratified analyses and the analysis of brain morphology on cognitive ability are provided in Tables 4, 8, 9 and 11. However, our results were not interpreted with a focus on p-values, but rather by examining causal effect estimates, the precision with which they were estimated (95% confidence intervals (CIs)), and on patterns of causal effects across cohorts and brain structures ^33, 34^.

### Effects of cognitive ability on brain morphology

#### Cortical thickness

In childhood, a greater cognitive ability was associated with a thicker postcentral cortex (β: 0.14; 95% CI: 0.04, 0.24) in the parietal lobe. In early adulthood, a one SD increase in cognitive ability was associated with a decreased thickness of the superior parietal and postcentral cortices of the parietal lobe (β: -0.23; 95% CI: -0.40, -0.03 and β: -0.25; 95% CI: -0.40, -0.03, respectively). In contrast, greater cognitive ability in early adulthood was associated with a greater thickness of the inferior temporal cortex (β: 0.26; 95% CI: 0.02, 0.50). For participants aged 45-68 years, there was evidence to suggest that cognitive ability was associated with a reduced thickness of the precuneus and insula (β: 0.11; 95% CI: 0.02, 0.22). Furthermore, there was evidence that cognitive ability was associated with a greater thickness of the lingual cortex of the occipital lobe, as well as that of the transverse temporal cortex for participants aged 45-60 years and 68-81 years, respectively (Figures 1a and 1b). Furthermore, a higher cognitive ability was associated with lower thickness in the medial orbitofrontal cortex in participants of ages 45-60 years and 60-68 years (β: -0.12; 95% CI: -0.22, -0.02 in participants of ages 60-68 years). A higher cognitive ability was also associated with a lower thickness of the pars opercularis and rostral middle frontal cortices in the participants of ages 60-68 years and the thickness of the rostral anterior cingulate cortex in the participants of ages 68-81 years. Additionally, a higher cognitive ability was associated with a reduced thickness of the superior parietal cortex in participants aged 68-81 years (β: -0.10; 95% CI: -0.18, -0.02). As observed in the participants aged 45-60 years, a greater cognitive ability was associated with a greater thickness of the transverse temporal cortex. Similar to participants aged 60-68 years, a greater cognitive ability was associated with a greater thickness of the superior temporal cortex (Figure 1b).

#### Cortical surface area

A higher cognitive ability was associated with a larger lateral orbitofrontal surface area in childhood (β: 0.09; 95% CI: 0.01, 0.18). This effect was also consistent in early adulthood (β: 0.22; 95% CI: 0.05, 0.39) (Figure 2a). In the temporal lobe, a one SD increase in cognitive ability was associated with a larger surface area of the banks of the superior temporal sulcus, entorhinal and inferior temporal cortices in participants aged 45-60 years (Figure 2b). Additionally, a higher cognitive ability was associated with a greater surface area of the caudal anterior cingulate in participants aged 60-68 years. Furthermore, a higher cognitive ability was associated with a smaller surface area of regions in the parietal lobe in mid-to-late adulthood, such as a smaller surface area of the cuneus and the superior parietal cortex in participants of ages 60-68 years and those aged 68-81 years, respectively (Figures 2a and Figure 2b). The largest and most consistent effect across all age cohorts was that of higher cognitive ability on a larger total cortical surface area (Figure 2b).

**Figure 2a.**
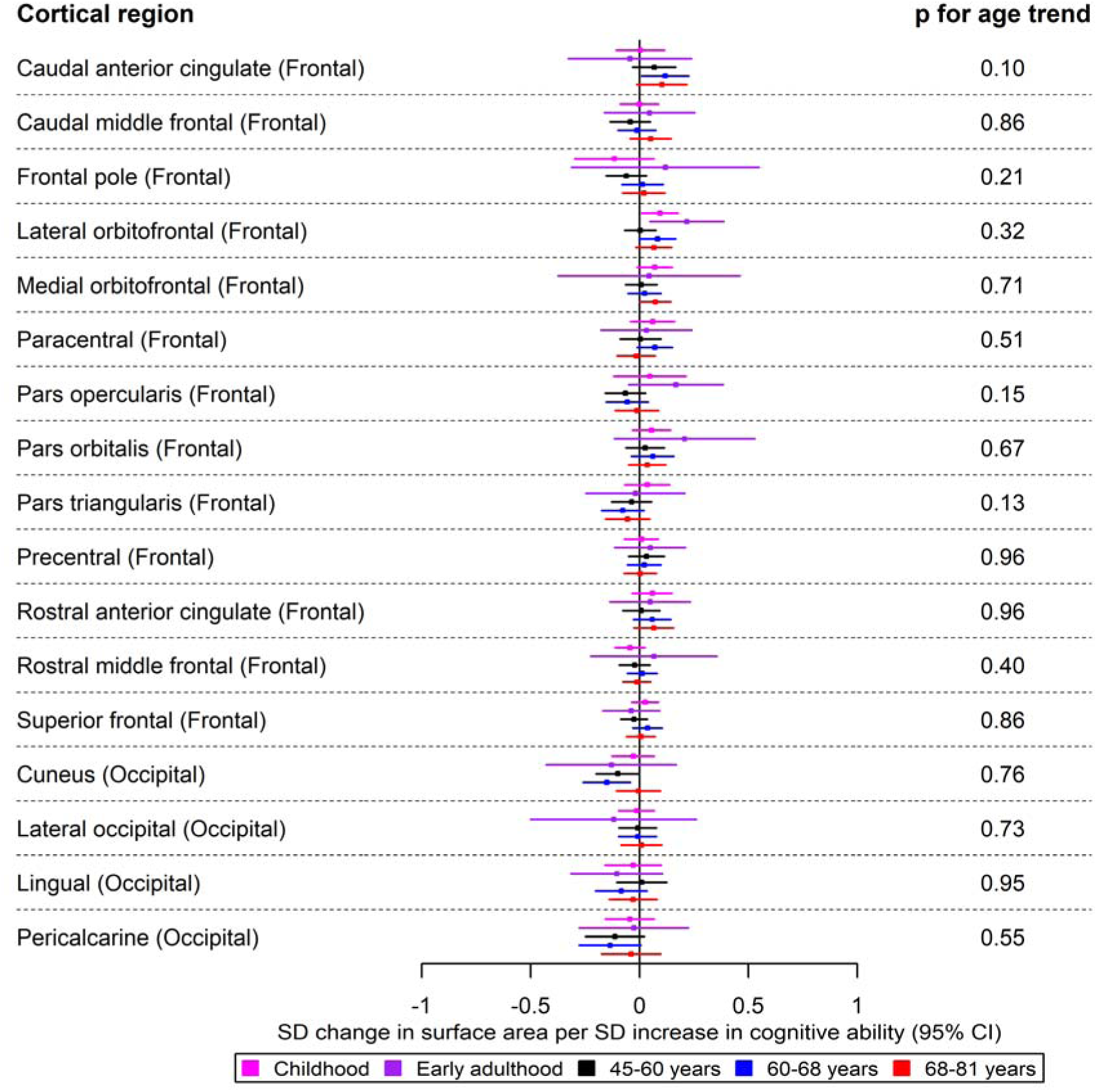
The causal effects of genetically predicted cognitive ability on the surface area of the frontal and occipital cortices at different ages across the life course (see Figure 2b for structures in the occipital, parietal, and temporal cortices, as well as total surface area). The childhood cohorts include meta-analysed effects of three peri-pubertal cohorts: ABCD, GEN R and IMAGEN. The early adulthood cohort includes meta-analysed effects of ALSPAC and IMAGEN (second wave for data collection), and the later adulthood cohort includes UK Biobank. Effect estimates represent SD changes in surface area. Regional measures were adjusted for total surface area. Where an effect estimate is missing, that structural measure was unavailable in that cohort.

**Figure 2b.**
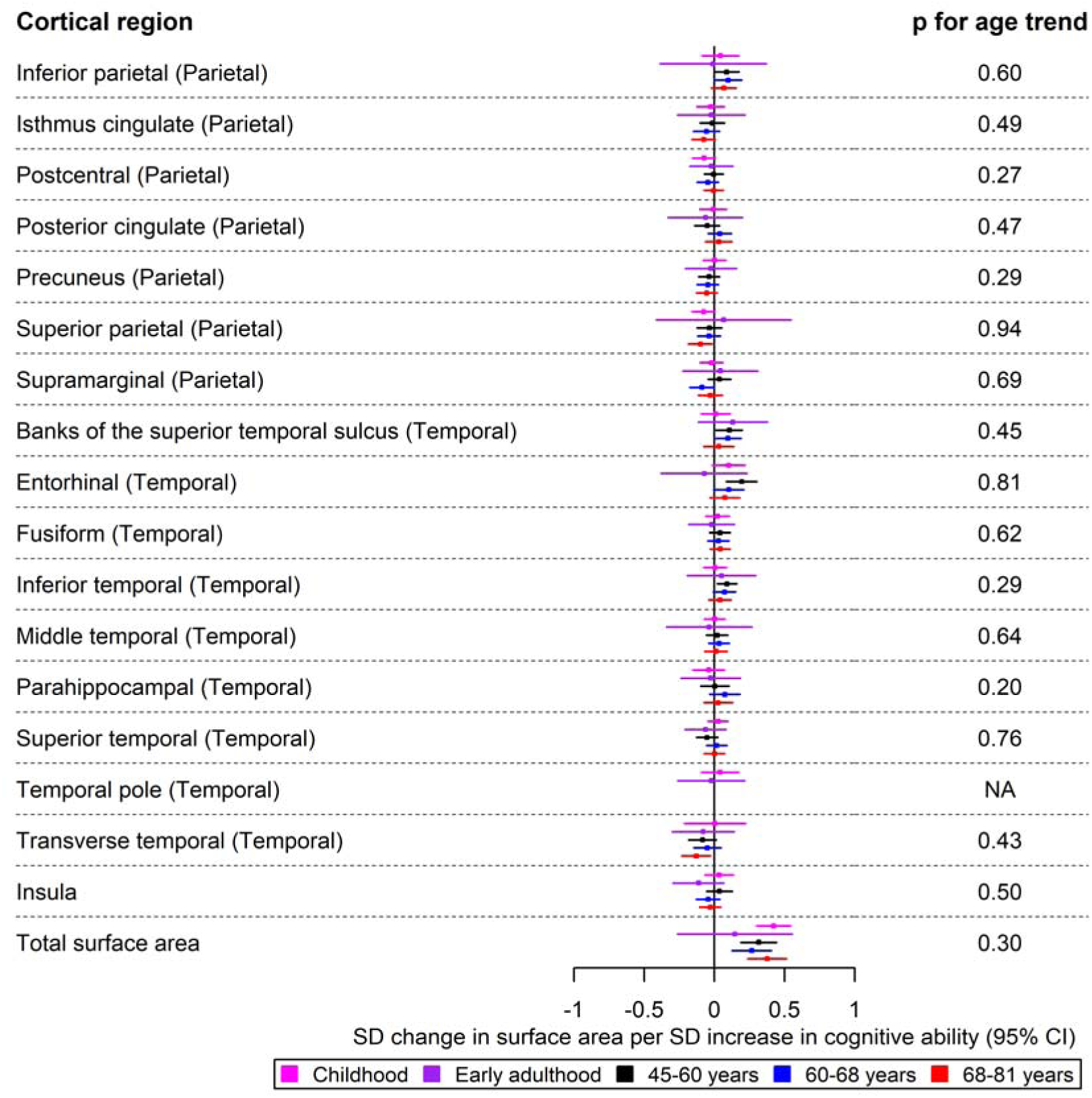
The causal effects of genetically predicted cognitive ability on the surface area of the parietal and temporal cortices at different ages across the life course. The childhood cohorts include meta-analysed effects of three peri-pubertal cohorts: ABCD, GEN R and IMAGEN. The early adulthood cohort includes meta-analysed effects of ALSPAC and IMAGEN (second wave for data collection), and the later adulthood cohort includes UK Biobank. Effect estimates represent SD changes in surface area. Regional measures were adjusted for total surface area. Where an effect estimate is missing, that structural measure was unavailable in that cohort.

#### Subcortical volumes

In childhood, higher cognitive ability was associated with differences in the volume of the accumbens (β: 0.28; 95% CI: 0.15, 0.41), brainstem (β: 0.13; 95% CI: 0.02, 0.23), hippocampal volume (β: 0.13; 95% CI: 0.01, 0.25) and total white matter volume (β: 0.09; 95% CI: 0.01, 0.18) (Figure 3). Additionally, a greater cognitive ability was associated with a larger volume of the accumbens in participants aged 60-68 years (β: 0.12; 95% CI: 0.01, 0.22) (Figure 3), and brainstem in participants of ages 68-81 years (Figure 3). The largest effect of cognitive ability was observed on estimated total intracranial volume in most cohorts (β: 0.40 per one SD increase in cognitive ability; 95% CI: 0.28, 0.52 in childhood); however, the evidence in the early adulthood cohort was weaker (β: 0.18 per one SD increase in cognitive ability; 95% CI: -0.03, 0.39) (Figure 3).

**Figure 3.**
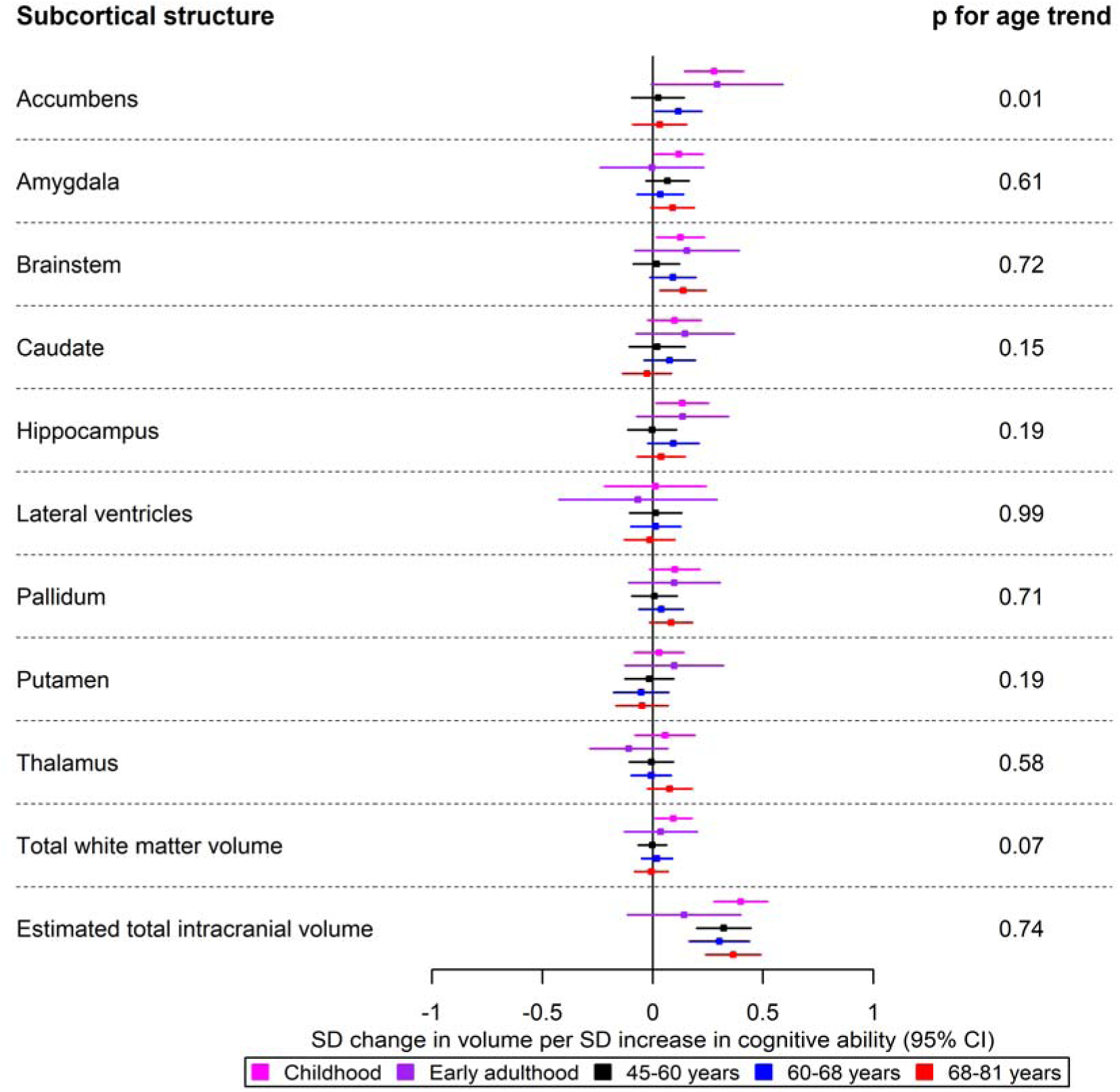
The causal effects of genetically predicted cognitive ability on the volume of subcortical structures at different ages across the life course. The childhood cohorts include meta-analysed effects of three peri-pubertal cohorts: ABCD, GEN R and IMAGEN. The early adulthood cohort includes meta-analysed effects of ALSPAC and IMAGEN (second wave for data collection), and the later adulthood cohort includes UK Biobank. Effect estimates represent SD changes in surface area. Regional measures were adjusted for the estimated total intracranial volume. Where an effect estimate is missing, that structural measure was unavailable in that cohort.

**Figure 4.**
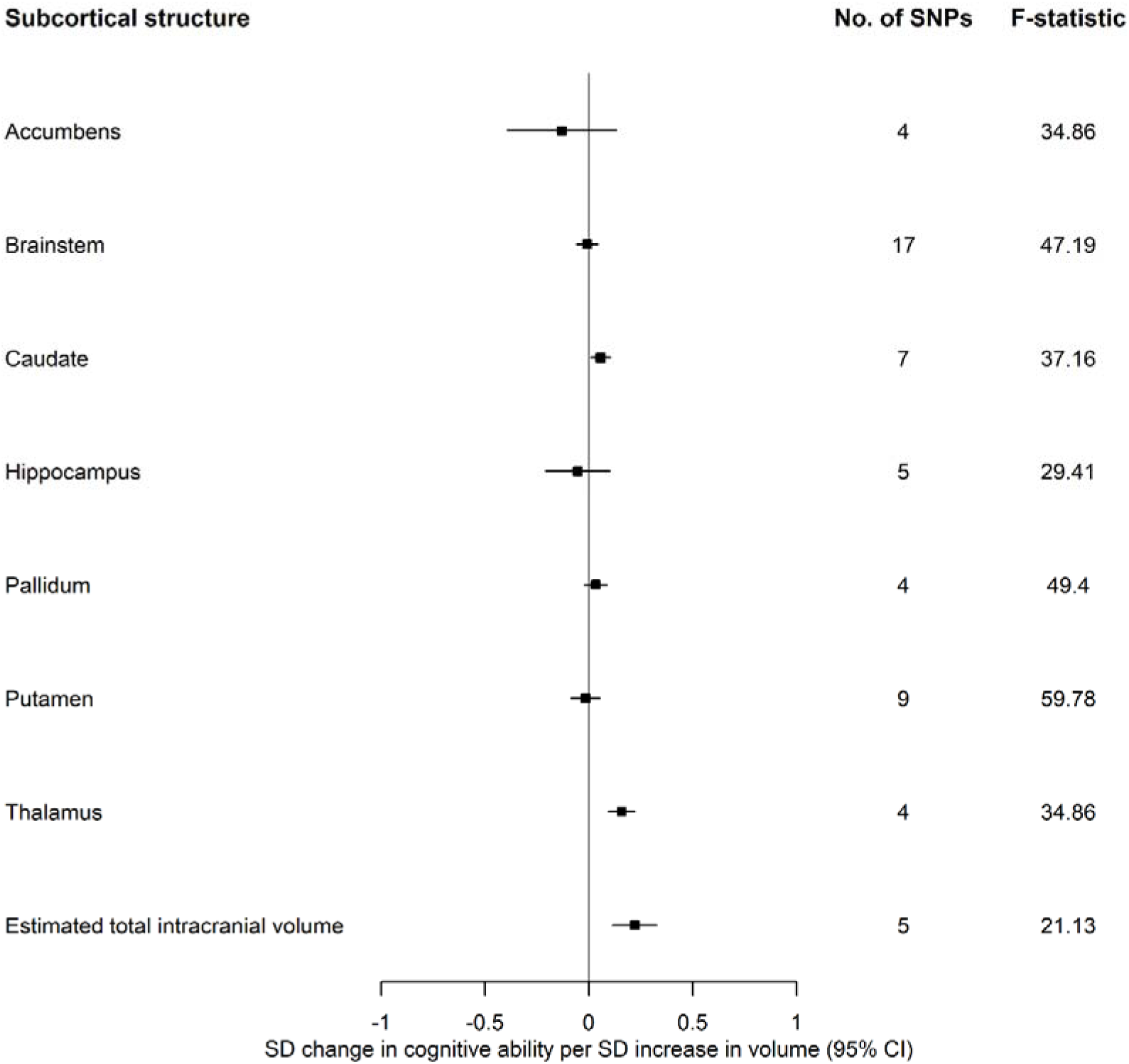
The causal effects of genetically predicted volume of subcortical structures from the ENIGMA consortium on cognitive ability.

### Replication analysis of cognitive ability on brain morphology using the ENIGMA consortium

Again, we employed two-sample Mendelian randomization to examine the effect of cognitive ability on brain morphology with the aim of replicating the findings from individual-level cohorts, using the ENIGMA consortium ^30, 35, 36^. As in the UK Biobank age-stratified analyses, a greater cognitive ability was associated with a lower thickness of the rostral anterior cingulate cortex and a greater thickness of the lingual cortex in the frontal lobe (Figures 1.1 and 1.2, Supplementary Material). A greater cognitive ability was associated with a lower thickness of the isthmus cingulate cortex in the parietal lobe, which is an effect not observed in the individual-level cohorts studied (Figure 1.1, Supplementary Material). Contrary to the early adulthood cohorts, where there is evidence of a small effect of cognitive ability on a greater thickness in the inferior temporal cortex, the opposite is observed with the ENIGMA consortium data. Furthermore, a higher cognitive ability was associated with a larger surface area of the banks of the superior temporal sulcus and the entorhinal cortex, while it was associated with a reduced surface area of the cuneus, precuneus and the superior parietal cortex (Figures 2.1 and 2.2, Supplementary Material). Except for the cuneus and precuneus which were not identified with the individual-level cohorts, all the other regions previously mentioned were observed in the mid-to-late adultood age-stratified analyses of surface area. As with those results, the largest effect was observed for total surface area (Figure 2.2, Supplementary Material). Similar to the age-stratified mid-to-late adulthood analyses, cognitive ability was associated with a larger volume of the accumbens, brainstem and estimated total intracranial volume (Figure 3, Supplementary Material). However, cognitive ability was also associated with increases in the the volume of the thalamus (β: 0.10; 95% CI: 0.02, 0.18) and amygdala (β: 0.13, 95% CI: 0.06, 0.21) in ENIGMA (Figure 3, Supplementary Material).

### Effects of brain morphology on cognitive ability

#### Cortical thickness

A greater thickness in the posterior cingulate region of the cortex was associated with a higher cognitive ability (β: 0.11; 95% CI: 0.03, 0.20) (Figure 4.1, Supplementary Material). Contrary, a greater thickness in the temporal pole was associated with a lower cognitive ability (β: -0.17; 95% CI: -0.26, -0.07) (Figure 4.2, Supplementary Material).

#### Cortical surface area

A larger surface area in the posterior cingulate cortex was associated with differences in cognitive ability (β: -0.10; 95% CI: -0.18, -0.02). A one SD larger surface area of the caudal middle frontal cortex was associated with a higher cognitive ability by 0.12 SD (95% CI: 0.07, 0.16) (Figure 5.1, Supplementary Material). A greater surface area of the temporal pole, instrumented by only 1 SNP (precluding investigation of potential bias due to horizontal pleiotropy) was associated with a greater cognitive ability (β: 0.46; 0.33, 0.59) (Figure 5.2, Supplementary Material). Furthermore, a larger total surface area was associated with a higher cognitive ability (β: 0.18; 95% CI: 0.07, 0.28) (Figure 5.2, Supplementary Material).

#### Subcortical structures

A larger thalamus (β: 0.16; 95% CI: 0.09, 0.22), caudate (β: 0.06; 95% CI: 0.01, 0.10) and estimated total intracranial volume (β: 0.22; 95% CI: 0.11,0.33) were associated with a greater cognitive ability (Figure 5).

**Figure 5.**
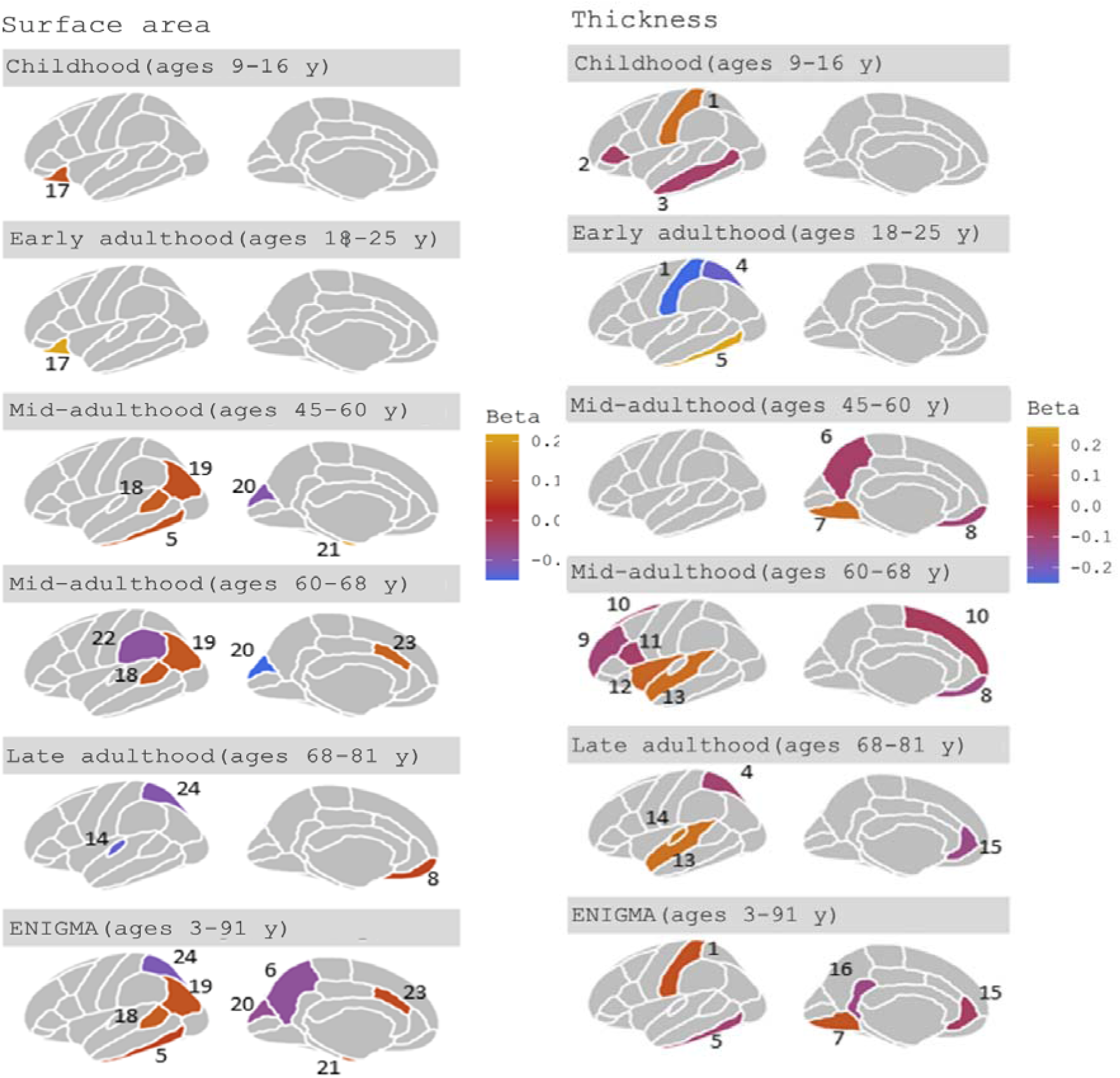
Regions affected by genetically predicted cognitive ability at different life stages at p<0.05. Below, the overall function of the regions in both hemispheres are described (in this study, we averaged measures across hemispheres). In this figure, we depict the regions from a lateral and medial view of the left hemisphere of the brain. The regions are: (1) the postcentral gyrus contains the primary somatosensory cortex, which receives sensory information about touch, temperature, pain and pressure from the contralateral side **^97^**; (2) the pars triangularis is part of the Broca’s area and plays a role in retrieving and selecting lexical and syntactic information from stimuli **^98, 99^**; (3) the middle temporal is involved in processing words and meaningful actions **^100^**; (4) the superior parietal cortex plays a role in features of attention and visuospatial perception, as well as manipulating and rearranging information **^43^**; (5) the inferior temporal gyrus is involved in visual object recognition **^101^**; (6) the precuneus has a role in a range of highly integrated tasks, such as navigation, episodic memory retrieval and self-reflection processes **^102^**; (7) the lingual gyrus plays a role in word processing **^103^**; (8) the medial orbitofrontal cortex is involved in goal-directed decision making **^104^**; (9) the rostral middle frontal gyrus is important for execution functions, such as working memory and emotional regulation and a study has shown that a higher thickness in the region is associated with stress-related cognitive bias which may encourage vulnerability to depression **^105, 106^**; (10) the superior frontal gyrus is associated with higher cognitive processes, particularly working memory **^107^**; (11) the pars opercularis, forms the Broca’s area, alongside the pars triangularis region and represents the interface between sensory stimuli and cognitive demands with motor representations of hand and face-associated actions **^108^**; (12) the insula is considered an ‘integral brain hub’ which connects a range of different functional systems associated with ‘sensory, emotional, motivational and cognitive processing’. The insula monitors the current environment, in addition to emotional and bodily states, and on the basis of experience, predicts how potential actions may affect survival and wellbeing **^109^**; (13) the superior temporal sulcus contains the auditory association cortex (Wernicke’s area) and is a multi-sensory integration site which plays a role in spoken word recognition and processing **^110^**; (14) the transverse temporal cortex has a role in early processing associated with understanding of speech (e.g., frequency and duration of sound). Language competence is believed to develop from decoding of auditory stimuli. However, the activation of the area is influenced from other modalities (e.g., observing faces producing speech without the associated auditory speech activates the transverse temporal gyrus in normal hearing **^111, 112^** and in deaf participants **^111, 112^**; (15) rostral anterior cingulate is located between limbic and cortical structures and is involved in processing emotion **^113, 114^**; (16) the isthmus of the cingulate gyrus has a less known role but there is evidence in its involvement in processing memory and pain, as well as mood symptoms such as anhedonia **^115^**; (17) the lateral orbitofrontal cortex integrates previous information with current information, in the anticipation of upcoming stimuli/events **^37^**; (18) the banks of the superior temporal sulcus are involved in speech and language processing, but its functional association with auditory areas is not well known **^116, 117^**; (19) the inferior parietal cortex plays a role in the processing and identification of visual stimuli and in memory and memory recollection to identify objects **^118^**; (20) the cuneus plays a role in basic visual processing and has been found to be associated with inhibitory control in individuals with bipolar disease **^119^**; (21) the entorhinal cortex acts as an interface between the hippocampus and the neocortex and thus, plays an important role in initial memory acquisition and retrieval (397), as well as in the processing of information relating to time **^40^** and spatial navigation (399); (22) the supramarginal gyrus has a role in phonological processing in language and memory tasks (400) in addition to overcoming egocentric bias to make judgments in social situations **^121^**; (23) the caudal anterior cingulate gyrus is involved in processing “sensory, motor, cognitive and emotional information” and affects the activity in other brain regions and changes “cognitive, motor, endocrine and visceral responses” **^122^**; (24) the superior parietal gyrus is thought to play a role in aspects of visuospatial perception and attention **^42^** and the manipulation and rearrangement of information in working memory **^43^**.

### Sensitivity analyses

#### Cognitive ability on brain morphology (age-stratified)

Various sensitivity analyses were performed to test for potential violations of key Mendelian randomization assumptions. The effect estimates using pleiotropy-robust estimators were directionally consistent with the IVW for most measures in all cohorts. However, there was some evidence of heterogeneity and pleiotropy in the effects of cognitive ability on cortical thickness for the medial orbitofrontal, rostral middle frontal, and postcentral areas of the cortex.

There was heterogeneity in the causal estimates of cognitive ability on the surface area of the caudal anterior cingulate cortex (Q=198, p=0.007), total surface area for participants of all age groups in the mid-to-late adulthood UK Biobank cohort (Q=315, p=2.15x10^-13^ for participants ages 45 to 60 years), well as surface area of the cuneus in the two older age tertiles (Q=211, P=0.001 for participants ages 45 to 60 years). Evidence of pleiotropic effects was identified only for cognitive ability SNPs on the total surface area for participants aged 60-68 years (p=0.002).

The directionality tests suggested the causal direction was from cognitive ability to all the brain structures examined (Table 1 of Sensitivity analyses in online repository). However, there was evidence that the SNPs instrumenting cognitive ability explained more variance in total surface area and estimated total intracranial volume (i.e., the outcome) than in cognitive ability (the exposure). When the SNPs that were in the wrong causal direction were removed for total surface area and estimated total intracranial volume (i.e., explained more variance in the outcome than exposure), the associations attenuated (e.g., total surface area IVW_tertile1_=0.07, 95% CI: -0.04, 0.18, Table 1 of Supplementary Material).

### Cognitive ability on brain morphology (replication)

There was heterogeneity for all identified structures except for lingual thickness and entorhinal surface area (Q statistic: 174.77, p=0.08). There was evidence of small pleiotropic effects for the thickness of the isthmus cingulate (p=0.04) and the surface area of the superior parietal (p=0.03) and whole cortex (p=0.03). The directionality tests suggested that the causal direction was from cognitive ability to cortical thickness, cortical surface area and subcortical structures in the ENIGMA consortium (i.e., SNPs for cognitive ability explained more variance in cognitive ability than in brain morphology) for all associated outcomes. Although the Steiger tests suggested the causal direction was false for estimated total intracranial volume in the age-stratified analyses, the test were equivocal (R^2^ in cognitive ability = 2.71%, R^2^ in estimated total intracranial volume=2.31%, p=0.19) in the ENIGMA analysis. Again, as opposed to the age-stratified UK Biobank analysis, the Steiger test suggested that the Mendelian randomization assumptions held for total surface area (R^2^ in cognitive ability=2.6%, R^2^ in total surface area=1.95%, Steiger test p=1.75x10^-4^).

### Brain morphology on cognitive ability

Due to an insufficient number of genetic instruments, the presence of pleiotropy for the thickness of the posterior cingulate and temporal pole on cognitive ability could not be tested, nor the causal effect estimates from any of the pleiotropy-robust methods. There was little evidence of pleiotropy (Egger intercept=-0.01, p=0.49) and heterogeneity (Q=1, p=0.53) in the causal effects of pericalcarine thickness on cognitive ability. For cortical surface area and subcortical structures, the effect estimates from IVW were consistent across all the pleiotropy-robust methods for all the identified outcomes and there was little evidence of pleiotropy and heterogeneity for most outcomes. Of the structures identified to influence cognitive ability, heterogeneity was detected in the causal estimates for the SNPs proxying the accumbens (N_SNP_=3, Q= 59, p=1.13x10^-12^) and estimated total intracranial volume (N_SNP_=4, Q=23, p=1.12x10^-4^). Still, there was little evidence of pleiotropy (p>0.05). The directionality tests suggested that the causal direction was from brain morphology to cognitive ability (i.e., SNPs for brain morphology explained more variance in brain morphology than in cognitive ability) for all associated outcomes.

## Discussion

This study examined the bidirectional effects between cognitive ability and cortical surface area and thickness, as well as subcortical brain morphology. There was little evidence of cognitive ability having large regional or age-dependent effects on the brain. However, there was consistent evidence of effects between cognitive ability and global measures of brain structure, in both directions.

### Effects of cognitive ability on structural brain morphology

Our results suggest cognitive ability affected the surface area and thickness of regions in the frontal, occipital, parietal, and temporal lobes, with the earliest effects observed on the lateral orbitofrontal cortex (in childhood and early adulthood). The lateral orbitofrontal cortex has been found to integrate previous information with current information in the anticipation of upcoming stimuli/events ^37^. A higher cognitive ability increased the surface area of the inferior temporal, entorhinal, and banks of the superior temporal sulcus and decreased the surface area of the transverse temporal cortex in age-stratified UK Biobank analyses. The temporal lobe has distinct functions such as perceptual processing of auditory stimuli, including speech, performing tasks requiring visual object discrimination and recognition, processing emotions, semantic knowledge, initial memory acquisition and retrieval ^38, 39^. Other important roles of the temporal lobe include processing information relating to time ^40^ and spatial navigation ^41^. In the parietal lobe, using age-stratified UK Biobank, the meta-analysed early adulthood cohort and the summary-level ENIGMA data, we found that a higher cognitive ability was associated with a lower cortical thickness and surface area of the superior parietal cortex. The superior parietal cortex is thought to play a role in aspects of visuospatial perception and attention ^42^, as well as the manipulation and rearrangement of information in working memory ^43^. In the occipital lobe, we found evidence to suggest that higher cognitive ability was associated with a thicker lingual cortex, which is involved in higher-order processing of emotional expression ^44^ and motional information ^45^. We observed that cognitive ability had the largest effects on total surface area and estimated total intracranial volume, but not mean thickness. This is in agreement with the findings of Mitchell and colleagues ^46^, showing the same associations with educational attainment and global measures of brain morphology. However, they found that educational attainment explained more variation in the surface area and cortical thickness of regions in the frontal and temporal lobes over-and-above the global effect. Although educational attainment and cognitive ability are similar, educational attainment is likely to also reflect non-cognitive skills, which may, at least in part explain the discrepancies in our findings.

Finally, individual differences in cognitive ability are associated with differences in the volume of the accumbens (childhood cohort meta-analysis) and brainstem (childhood cohort meta-analysis and adults of ages 68-81 years). A previous analysis in the IMAGEN cohort showed that cognitive ability is associated with an increase in the grey matter volume of the striatum (the accumbens is in the striatum) and functional activation in the accumbens ^47^, induced by reward-prediction error cues which is known to affect dopamine neurotransmission. Dopamine has an established role in cognitive ability and decision-making ^48–50^.

### Effects of structural brain morphology on cognitive ability

A larger caudate, thalamus and estimated total intracranial volume were associated with a higher cognitive ability. The effect of the thalamus on cognitive ability (β_IVW_=0.16), adjusted for estimated total intracranial volume, has a comparable strength of association to estimated total intracranial volume (β_IVW_=0.22) and is above and beyond what would be considered a global change. This may suggest that larger estimated total intracranial volume in participants with higher cognitive ability may be attributed to a larger thalamic volume. This observation aligns with a non-genetic study in the UK Biobank ^51^, wherein thalamic volume and white matter microstructure of thalamic and association fibres display the highest levels of correlation with a latent factor of general cognitive ability. Medial thalamic nuclei are densely interconnected with prefrontal and temporal cortices and control working memory ^52, 53^ and attentional control ^54^, while posterior nuclei project to occipital cortices and aid in visual processing. Functional and structural studies suggest that the thalamus contributes to the pathogenesis of diseases such as dementia ^55^, Parkinson’s disease ^56^, and schizophrenia ^57^. The caudate, another striatal structure with evidence of association with cognitive ability has afferent and efferent connections to the prefrontal and anterior cingulate cortices ^58^ and is highly innervated with dopamine neurons, which support brain networks for seeking, evaluation, value learning, orienting, cognition and general motivation ^59^.

For the cortical measures and in concordance with the P-FIT model ^11^, there is evidence that a thicker posterior cingulate cortex and a larger surface area of the caudal middle frontal cortex and temporal pole increase cognitive ability. Functionally, these regions have been shown to be associated with internally directed cognition (memory retrieval or planning for the future) ^60^, the control and the reorientation of attention in response to exogenous stimuli ^60, 61^. A larger cortical surface area can boost information processing by accommodating more cortical columns ^62, 63^, which are the functional units of the cortex ^64^. The larger number of cortical columns is expected to correspond to fewer intercolumnar connections, which is thought to enhance the functional specificity of cortical columns and reduce the overlap in their representations, consequently increasing their capacity to store information ^65, 66^. Contrarily, cortical thickness is linked to neuronal migration, the number neurons, dendritic arborisation, and the support provided by glial cells within cortical columns ^66^. Observations of decreases in thickness in relation to cognitive ability have been reported to reflect pruning of weak neural connections, resulting in a more organised neural network^66^. In our study, the genetic instruments for the brain morphology measures were extracted from a consortium which comprised mainly of adults. Hence, the mechanisms which mediate the relationship between cognitive ability and cortical thickness may be different across the lifespan (e.g., synaptic pruning in childhood and myelin loss in late adulthood).

### Age-varying effects of cognitive ability on brain morphology

Overall, there was little evidence of age-varying effects of cognitive ability on structural brain morphology; the positive effects persisted across the life course with a relatively similar magnitude. This suggests that a higher cognitive ability results in larger brain structures in early life and therefore, given a similar rate of neurodegeneration, people with higher cognitive ability will have, on average, larger brain structures in old age. Theories of ’brain reserve’ and ’brain maintenance’ are used to explain the brain’s ability to be resilient against processes of aging, neurological diseases, and cognitive decline. Brain maintenance describes the brain’s capacity to maintain neurochemical, structural and functional brain health over time, irrespective of ageing processes ^67^. Brain reserve refers to neuroanatomical resources such as a larger brain size or a greater number of neurons which increase the brain’s capacity to tolerate age-related changes or pathological process without displaying symptoms of neurological disease such as cognitive decline ^68^. The findings of our study support the former hypothesis, indicating that early brain morphology (i.e., brain reserve) is underpinned by early cognitive ability, potentially through neurodevelopmental mechanisms. In a longitudinal study (N=974; ages 4-88 years), trajectories of change in cortical surface area in individuals with higher and lower cognitive abilities followed parallel trajectories throughout their lifespan. Adaptive responses to the environment are thought to decrease with age, as stability becomes important in supporting social continuity and the energy requirement for adaptability is higher in older individuals due to accumulated damage, influenced by evolved limitations in bodily maintenance ^69^.

### Potential explanations for the observed relationships

The observed findings of more and larger effects of cognitive ability on structural brain morphology than vice versa may be counterintuitive, given the early developmental origin of structural brain morphology. A potential explanation for the directionality may be dynastic effects (and shared parent-offspring genetics for the two traits). For instance, in our analyses, there may be a confounding path linking SNPs for cognitive ability to brain morphology via the correlation between the offspring’s genes for cognitive ability (inherited from the parent) and the environment the parents create for their offspring. A different explanation may be due to pleiotropic effects, which can affect Mendelian randomization studies. However, several studies have shown experience-dependent structural neuroplasticity, where the brain structure changes in response to tasks ^70–72;^ albeit the timespan that these effects last for are not well-established. The exploration selection refinement (ESR) model of human brain plasticity, which is motivated from developmental theory and animal studies, suggests that when individuals learn new skills, the microcircuits in the brain are initially widely explored resulting in higher neural activity. This consequently induces structural changes in neurons ^73, 74^, such as new dendritic spines ^75^ and increased myelination ^76, 77^. This process is influenced by reinforcement learning and neurotransmitters (e.g., dopamine) ^78^. The exploration phase is followed by experience-dependent selection and refinement of reinforced microcircuits and the retraction of structures associated with unselected circuits ^69^. Given the MR results, it is not possible to disentangle how exactly these effects unfold.

## Strengths and limitations

Most genetic and observational studies focus on cognitive ability and anatomical features of the cortex. We examined both cortical and subcortical structures and showed that subcortical structures might play an even greater role in affecting differences in cognitive ability than anatomical features of the cortex. Additionally, genetic studies have examined genetic correlations between brain morphology and cognitive ability using measures of regional cortical volume. Cortical surface area and thickness are genetically and phenotypically independent ^17, 79^ and analyses of cortical volume (which reflects a combination of both cortical surface area and thickness), may not provide clear insight as to which one (if either) drives observed associations. Additionally, a life course approach was taken using five cohorts capturing different life stages to see if effects differ in the earlier neurodevelopmental or later neurodegenerative periods. The fewer regional effects of differences in cognitive ability on surface area in childhood/early adulthood compared to mid- and later adulthood despite the presence of effects on total surface area and estimated total intracranial volume, may be due to developmental noise ^80^ or non-linear trajectories^81^.

As mentioned previously, the two groups of phenotypes analysed are susceptible to bias due to dynastic effects. This study design is optimal for examining the question with the data currently available. However, once there is availability of a well-powered cohort with neuroimaging, genetic, and family data, within-family Mendelian randomization studies could minimise risk of confounding of genetic instruments by dynastic effects, by including a fixed effect for shared familial environmental effects. Additionally, our study included volumetric neuroimaging markers and consequently, inferences cannot be made for other neuroimaging markers, such as brain microstructure or functional connectivity. Finally, pleiotropy is a phenomenon which may affect Mendelian randomization study findings, where a SNP affects an outcome through pathways other than the exposure. However, our sensitivity analyses for most structures showed little evidence of pleiotropy, and where they did, we have indicated heterogeneity and pleiotropy statistics.

## Conclusion

Cognitive ability had effects on cortical surface area and estimated total intracranial volume from early in childhood and across the life course, suggesting that it may to be useful to find ways to improve cognitive ability to increase brain reserve and potential neuroprotective effects. Further research needs to integrate data from other modalities into structural studies, such as these, to establish the functional role of these differences in brain structure.

## Methods

### Data

#### Cognitive ability GWAS

Genetic instruments for cognitive ability were extracted from a Multi-Trait Analysis of GWAS (MTAG) of 269,867 European participants ^10^, where cognitive ability was measured through verbal-numerical (VNR) test scores. All the VNR scores were controlled for age, sex, assessment centre, genotype batch, array, and 40 principal components. The genome-wide significant SNPs (p<5x10^-8^) were re-clumped at an r2 threshold of <0.001 within a 10mb window using the 1000 genomes reference panel to ensure independence of SNPs ^82^. We identified 153 SNPs for cognitive ability. SNP coefficients reflect the SD increase in verbal-numeric reasoning test scores (SD=15 points) per allele increase.

#### Brain morphology GWAS

GWAS of MRI-derived neuroimaging measures of thickness and surface area of 34 regions defined by the Desikan-Killiany atlas ^83^ in five cohorts across the life course were used in our analyses. For the peri-pubertal period, Generation R (a prospective population-based birth cohort from Rotterdam, the Netherlands, N=1,175, age range 8.71 to 11.99) ^21, 84^, the Adolescent Brain Cognitive Development study (ABCD, N= 5,022, age range 8.92 to 11.00 at baseline) ^19, 20^ and IMAGEN ^25^, a multi-centre genetic neuroimaging study recruiting adolescents from secondary schools across Europe, N=1,739, age range = 12.94 to 16.04) were used. For early adulthood, the Avon Longitudinal Study of Parents and Children (ALSPAC) ^22–24, 85^ (N=776, age range 18.00 to 24.5 years), and the second wave of IMAGEN data collection (N=1,161, age range=17.68-21.53) were used. ALSPAC consists of data on offspring of pregnant women resident in Avon, UK with expected delivery dates in 1991/1992. The core sample includes 13,988 children but we used data from a subset of ALSPAC offspring invited to participate in three different neuroimaging sub-studies; the ALSPAC Testosterone study, the ALSPAC Psychotic Experiences (PE) study and the ALSPAC Schizophrenia Recall-by-Genotype Study ^85^. Please note that the study website contains details of all the data that is available through a fully searchable data dictionary and variable search tool (http://www.bristol.ac.uk/alspac/researchers/our-data/). UK Biobank was stratified into age-ordered tertiles to examine age-specific effects in adulthood. The UK Biobank is a population-based study of 503,325 participants who were recruited from across Great Britain between 2006 and 2010 ^86^ (N=9,377 per tertile, youngest age tertile = 45 to 60 years, middle age tertile = 60 to 68 years and oldest age tertile = 68 to 81 years). We also used volume measures of nine subcortical structures and total white matter, as well as the global measures of mean thickness, total surface area and estimated total intracranial volume. Finally, we used summary data for the same structural brain measures as in the individual-level data cohorts from the ENIGMA consortium GWAS (N for subcortical structures=37,741; N for estimated total intracranial volume and hippocampus; N for cortical regions=33,392), which included study samples from various studies, approximately 75% of which are population-based ^30–32, 35, 36^. SNPs for estimated total intracranial volume and hippocampal volume were identified from GWAS by Adams et al ^31^ and Hibar et al ^32^, respectively, but we used the effect sizes from an earlier GWAS ^36^ due to data restrictions in investigations relating to cognitive ability-associated genetics in the CHARGE summary statistics. For the MR examining the effects of brain structure on cognitive ability, genetic variants for brain structure were obtained from the ENIGMA consortium GWAS. All GWAS for regional cortical thickness, surface area and subcortical volumes in our analyses were adjusted for global mean cortical thickness, total surface area and estimated total intracranial volume, respectively, to identify region-specific effects.

### Statistical Analyses

#### Estimating the causal effects of cognitive ability on brain morphology

MR is a form of instrumental variable analysis, which uses SNPs to proxy for environmental exposures to estimate the causal effects of an exposure on an outcome ^26^. Two-sample MR is where the association of the genetic variant and the exposure and outcome are obtained from separate GWAS and this method was used for all the analyses in this study. For MR to generate unbiased causal effect estimates, each genetic variant that is used as an instrumental variable must satisfy three assumptions: (1) that it is associated with the exposure (relevance assumption), (2) that it is not associated with the outcome through a confounding pathway (exchangeability assumption), and (3) is only associated with the outcome through the exposure (exclusion restriction assumption). More details on terms related to MR can be found in the MR dictionary ^87^. SNPs associated with cognitive ability were extracted from each brain structure GWAS at p≤5×10^-8^. Where a SNP for cognitive ability was not available in the brain structure GWAS, proxy SNPs identified at r2>0.80 were used across all individual-level datasets (SNPs rs1174546, rs17381294, rs1105307, rs10760199, rs8028238, rs4982712 proxied rs28420834, rs61787263, rs7033137, rs4446794, rs55894132, rs12900061, respectively). The cognitive ability GWAS were harmonised with the brain structure GWAS in IMAGEN, Generation R, ABCD, ALSPAC and the UK Biobank. Random-effects IVW regression, which assumes no directional horizontal pleiotropy was employed in the analyses ^88^. The F-statistic was used as a measure of instrument strength ^89^. We meta-analysed the effects of SNPs for cognitive ability on structural brain measures for the three peri-pubertal cohorts, using random-effects models. Additionally, to test whether there is strong evidence of an age-varying effect, the metareg command in STATA ^90^ was used to obtain a p-value for the difference in the effects observed between childhood, early adulthood, and the different stages of adulthood.

#### Estimating the causal effect of brain structures on cognitive ability

Using the ENIGMA consortium ^30–32, 35^, we extracted SNPs associated with structural brain measures at 5x10^-8^. SNPs were clumped using r2>0.001 and a physical distance for clumping of 10,000 kb. Analyses were performed as described previously. We identified considerably less genetic instruments for brain structure than cognitive ability (N_min_=1, N_max_=17)

### Sensitivity analyses

IVW regression assumes no directional horizontal pleiotropy and only provides unbiased causal effect estimates when there is balanced or no horizontal pleiotropy. IVW estimates were compared to those from Egger regression ^91^, weighted median ^92^ and weighted mode ^93^ which relax this assumption. Heterogeneity in the causal estimates (which can indicate pleiotropy) was calculated using Cochran’s Q statistic ^91^. For meta-analyses, these heterogeneity statistics were examined in each cohort and are available in the online repository. Additionally, to exclude the possibility that the genetic variants used as proxies for cognitive ability are better instruments for brain structures and vice versa (i.e., to test that the hypothesized causal direction is correct for each SNP used), directionality (Steiger) tests were used ^94^. Steiger tests were not performed for the analyses in childhood and early adulthood as Steiger filtering has been shown to be biased in small samples ^95^. Where the hypothesized direction was false, SNPs explaining greater variance in the outcome than the exposure were removed, to examine change in the estimated causal effects.

### Replication

As with the previous analysis, two-sample MR was used to examine the effects of structural brain morphology derived from the ENIGMA consortium on cognitive ability. There is overlap between ENIGMA and some of the individual-level cohorts. However, it has been shown that sample overlap results in little bias in the presence of strong instruments (i.e., F>10) ^96^.

### Data availability

The ENIGMA consortium MRI summary measures from genetic association analyses of estimated total intracranial volume, subcortical structures, as well as cortical thickness were requested online. The ABCD Study data are openly available to qualified researchers for free (https://nda.nih.gov/abcd/request-access). Requests for Generation R data should be directed toward the management team of the Generation R Study, which has a protocol of approving data requests. For access to IMAGEN data, researchers may submit a request to the IMAGEN consortium (https://imagen-europe.com/resources/imagen-project-proposal/). ALSPAC details and data descriptions are available on their website (www.bristol.ac.uk/alspac/researchers/access), where applications for individual-level data can be made (managed access). UK Biobank data are available through a procedure described on their website (http://www.ukbiobank.ac.uk/using-the-resource/). UK Biobank is approved by the National Health Service National Research Ethics Service (ref 11/NW/0382; UK Biobank application number 48970. Ethics approval for the study was obtained from the ALSPAC Ethics and Law Committee and the Local Research Ethics Committees and informed consent for the use of data collected via questionnaires and clinics was obtained from participants. In Generation R, all study protocols and measurements assessed in each wave of data collection were approved by the Medical Ethical Committee (MEC 198.782/2001/31) of the Erasmus MC, University Medical Center Rotterdam. The IMAGEN study was approved by the institutional ethics committee of Kings College London, University of Nottingham, Trinity College Dublin, University of Heidelberg, Technische Universität Dresden, Commissariat á l Energie Atomique et aux Energies Alternatives, and University Medical Center at the University of Hamburg in accordance with the Declaration of Helsinki. The UCSD IRB approved all data collection protocols for ABCD. IRB number: 160091. All analyses in this study used de-identified data, therefore no additional IRB approval was required.

### Code availability

Code is available at https://github.com/rskl92/intelligence_brain_morphology_bidirectional_MR.

## Supporting information

Supplementary Results

## Data Availability

The ENIGMA consortium MRI summary measures from genetic association analyses of estimated total intracranial volume, subcortical structures, as well as cortical thickness were requested online. The ABCD Study data are openly available to qualified researchers for free (https://nda.nih.gov/abcd/request-access). Requests for Generation R data should be directed toward the management team of the Generation R Study, which has a protocol of approving data requests. For access to IMAGEN data, researchers may submit a request to the IMAGEN consortium (https://imagen-europe.com/resources/imagen-project-proposal/). ALSPAC details and data descriptions are available on their website (www.bristol.ac.uk/alspac/researchers/access), where applications for individual-level data can be made (managed access). UK Biobank data are available through a procedure described on their website (http://www.ukbiobank.ac.uk/using-the-resource/).

https://github.com/rskl92/intelligence_brain_morphology_bidirectional_MR

## Acknowledgments

Data used in the preparation of this article were obtained from the Adolescent Brain Cognitive Development (ABCD) Study, IMAGEN, ALSPAC, Generation R and UK Biobank. The ABCD study data (https://abcdstudy.org) are held in the NIMH Data Archive (NDA). This is a multisite, longitudinal study designed to recruit more than 10,000 children age 9-10 years and follow them over 10 years into early adulthood. The ABCD Study is supported by the National Institutes of Health and additional federal partners under award numbers U01DA041048, U01DA050989, U01DA051016, U01DA041022, U01DA051018, U01DA051037, U01DA050987, U01DA041174, U01DA041106, U01DA041117, U01DA041028, U01DA041134, U01DA050988, U01DA051039, U01DA041156, U01DA041025, U01DA041120, U01DA051038, U01DA041148, U01DA041093, U01DA041089, U24DA041123, U24DA041147. A full list of supporters is available at https://abcdstudy.org/federal-partners.html. A listing of participating sites and a complete listing of the study investigators can be found at https://abcdstudy.org/consortium_members/. ABCD consortium investigators designed and implemented the study and/or provided data but did not necessarily participate in analysis or writing of this report. This manuscript reflects the views of the authors and may not reflect the opinions or views of the NIH or ABCD consortium investigators. The ABCD data repository grows and changes over time. The ABCD data repository grows and changes over time. The ABCD data used in this report came from [NIMH Data Archive Digital Object Identifier (10.15154/1503209)]. We are extremely grateful to all the families who took part in this study, the midwives for their help in recruiting them, and the whole ALSPAC team, which includes interviewers, computer and laboratory technicians, clerical workers, research scientists, volunteers, managers, receptionists and nurses. The UK Medical Research Council and Wellcome (Grant ref: 217065/Z/19/Z) and the University of Bristol provide core support for ALSPAC. A comprehensive list of grants funding is available on the ALSPAC website (http://www.bristol.ac.uk/alspac/external/documents/grant-acknowledgements.pdf). The ALSPAC-Testosterone study was funded by the National Institutes of Health, U.S.A. (R01MH085772 to TP). The ALSPAC-PE study was funded by a grant from the UK Medical Research Council (G0901885). ASD was also supported by the National Institutes of Health Research Biomedical Research Centre at the South London & Maudsley Hospital Foundation NHS Trust and the IoPPN, King’s College London. The ALSPAC SCZ-RbG study was funded by grant MR/K004360/1 from the Medical Research Council (MRC) titled: “Behavioural and neurophysiological effects of schizophrenia risk genes: a multi-locus, pathway based approach” and by the MRC Centre for Neuropsychiatric Genetics and Genomics (G0800509) and the NIHR Bristol Biomedical Research Centre. GWAS data was generated by Sample Logistics and Genotyping Facilities at Wellcome Sanger Institute and LabCorp (Laboratory Corporation of America) using support from 23andMe. IMAGEN is supported by the European Union-funded FP6 Integrated Project IMAGEN (Reinforcement-related behaviour in normal brain function and psychopathology) (LSHM-CT- 2007- 037286), the Horizon 2020 funded ERC Advanced Grant ‘STRATIFY’ (Brain network based stratification of reinforcement-related disorders) (695313), Human Brain Project (HBP SGA 2, 785907, and HBP SGA 3, 945539), the Medical Research Council Grant ‘c-VEDA’ (Consortium on Vulnerability to Externalizing Disorders and Addictions) (MR/N000390/1), the National Institute of Health (NIH) (R01DA049238, A decentralized macro and micro gene-by-environment interaction analysis of substance use behavior and its brain biomarkers), the National Institute for Health Research (NIHR) Biomedical Research Centre at South London and Maudsley NHS Foundation Trust and King’s College London, the Bundesministeriumfür Bildung und Forschung (BMBF grants 01GS08152; 01EV0711; Forschungsnetz AERIAL 01EE1406A, 01EE1406B; Forschungsnetz IMACMind 01GL1745B), the Deutsche Forschungsgemeinschaft (DFG grants SM 80/7-2, SFB 940, TRR 265, NE 1383/14-1), the Medical Research Foundation and Medical Research Council (grants MR/R00465X/1 and MR/S020306/1), the National Institutes of Health (NIH) funded ENIGMA (grants 5U54EB020403-05 and 1R56AG058854-01). Further support was provided by grants from: – the ANR (ANR-12-SAMA-0004, AAPG2019 – GeBra), the Eranet Neuron (AF12-NEUR0008-01 – WM2NA; and ANR-18-NEUR00002-01 – ADORe), the Fondation de France (00081242), the Fondation pour la Recherche Médicale (DPA20140629802), the Mission Interministérielle de Lutte-contre-les-Drogueset-les-Conduites-Addictives (MILDECA), the Assistance-Publique-Hôpitaux-de-Paris and INSERM (interface grant), Paris Sud University IDEX 2012, the Fondation de l’Avenir (grant AP-RM-17-013), the Fédération pour la Recherche sur le Cerveau; the National Institutes of Health, Science Foundation Ireland (16/ERCD/3797), U.S.A. (Axon, Testosterone and Mental Health during Adolescence; RO1 MH085772-01A1), and by NIH Consortium grant U54 EB020403, supported by a cross-NIH alliance that funds Big Data to Knowledge Centres of Excellence. The general design of Generation R Study is funded by the Erasmus Medical Center, Rotterdam, ZonMw, the Netherlands Organization for Scientific Research (NWO), and the Ministry of Health, Welfare and Sport, and is conducted by the Erasmus Medical Center in close collaboration with the Faculty of Social Sciences of the Erasmus University Rotterdam, and the Stichting Trombosedienst & Artsenlaboratorium Rijnmond (STAR-MDC), Rotterdam. This project received funding from the European Union’s Horizon 2020 Research and Innovation Programme (EarlyCause; project no: 848158), the HorizonEurope Research and Innovation Programme (FAMILY, grant agreement No 101057529; HappyMums, grant agreement No 101057390) and the European Research Council (TEMPO; project no: 101039672). The UK Biobank data used in this work were obtained from UK Biobank Data Application 48970. We thank UK Biobank for making the data available, and to all UK Biobank study participants, who generously donated their time to make this resource possible.

## Contributions

RKL designed the study, cleaned, and performed data analyses, interpreted the results, wrote the first draft of the manuscript and revised the manuscript. RKL, ELA, NMD, YBS and LDH conceptualized the study; RKL performed the statistical analyses in the Avon Longitudinal Study of Parents and Children (ALSPAC), the Adolescent Brain Cognitive Development study (ABCD), IMAGEN, UK Biobank, and the Enhancing Neuroimaging Genetics through Meta-analysis (ENIGMA) consortium; IS performed data cleaning and statistical analysis in Generation R. RKL, IS, ELA, NMD, TW, CC, LDH, YBS, TP, TB, AH, FN discussed the results and provided comments on the manuscript.

