## Supplementary Results for "The bidirectional effects between cognitive ability and brain morphology: A life course Mendelian randomization analysis"

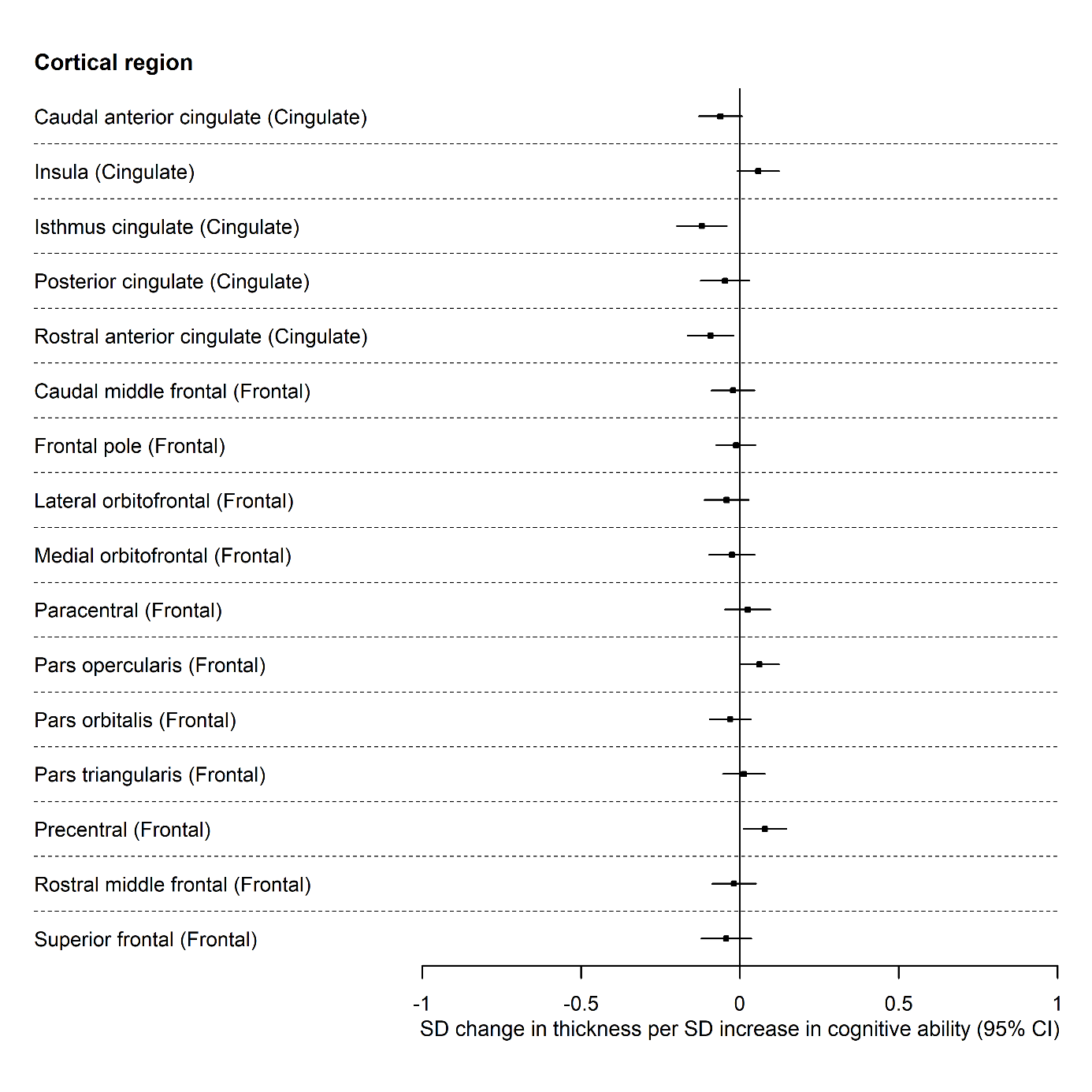

Figure 1.1 The causal effects of genetically predicted cognitive ability on the thickness of the cingulate, and frontal cortices from the ENIGMA consortium.

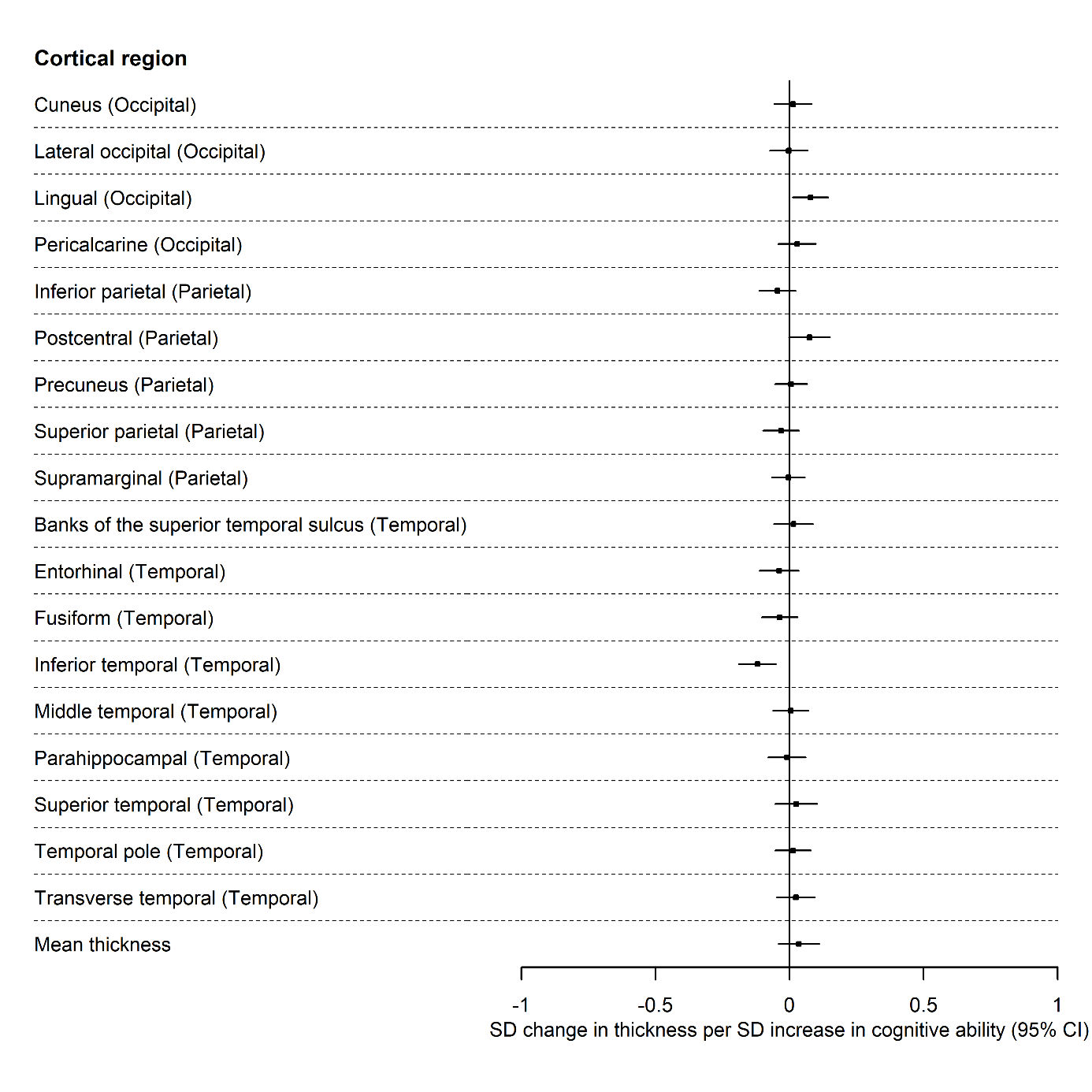

Figure 1.2 The causal effects of genetically predicted cognitive ability on the thickness of the occipital, parietal, and temporal cortices and mean thickness from the ENIGMA consortium.

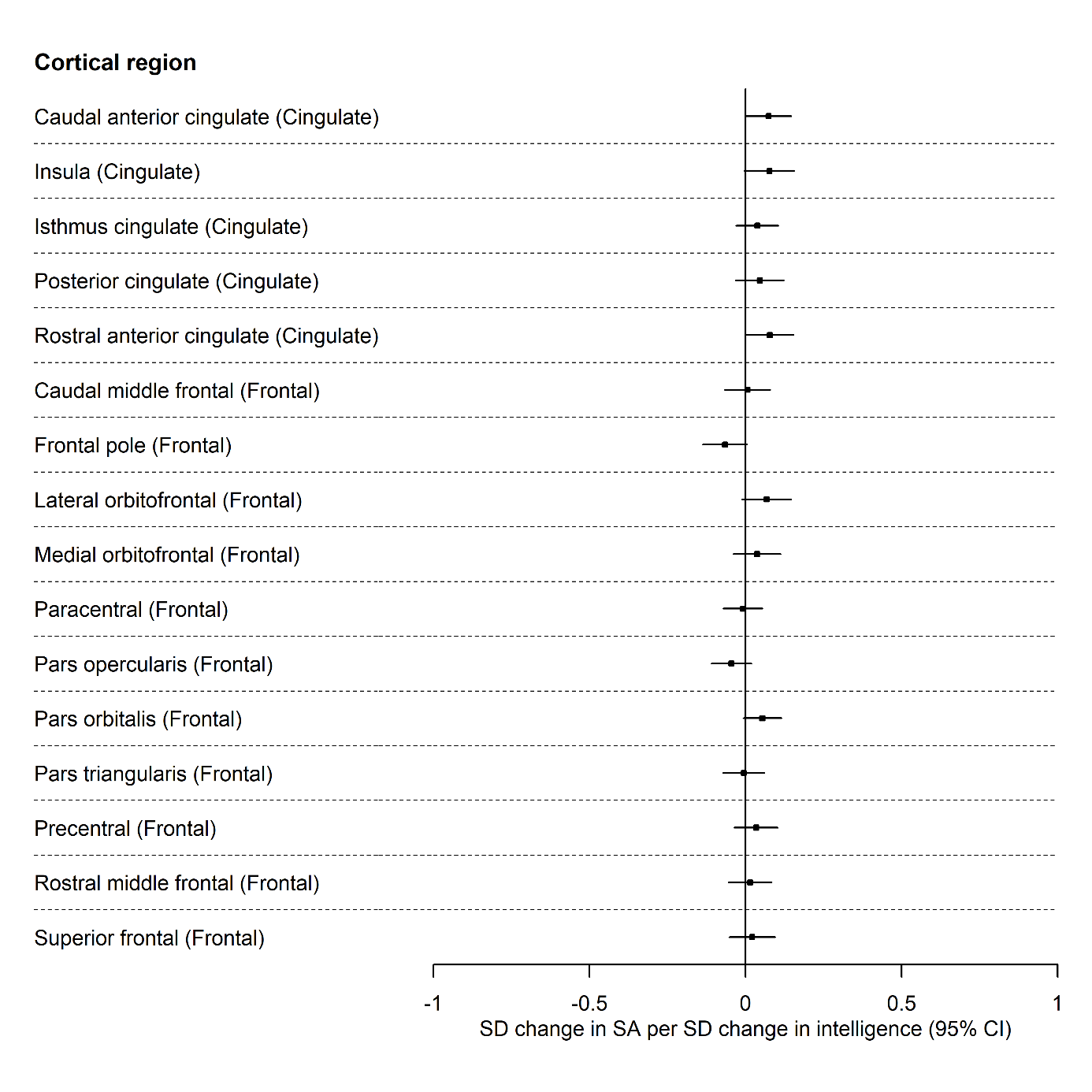
**Figure 2.1** The causal effects of genetically predicted cognitive ability on the surface area of the cingulate, and frontal cortices from the ENIGMA consortium.

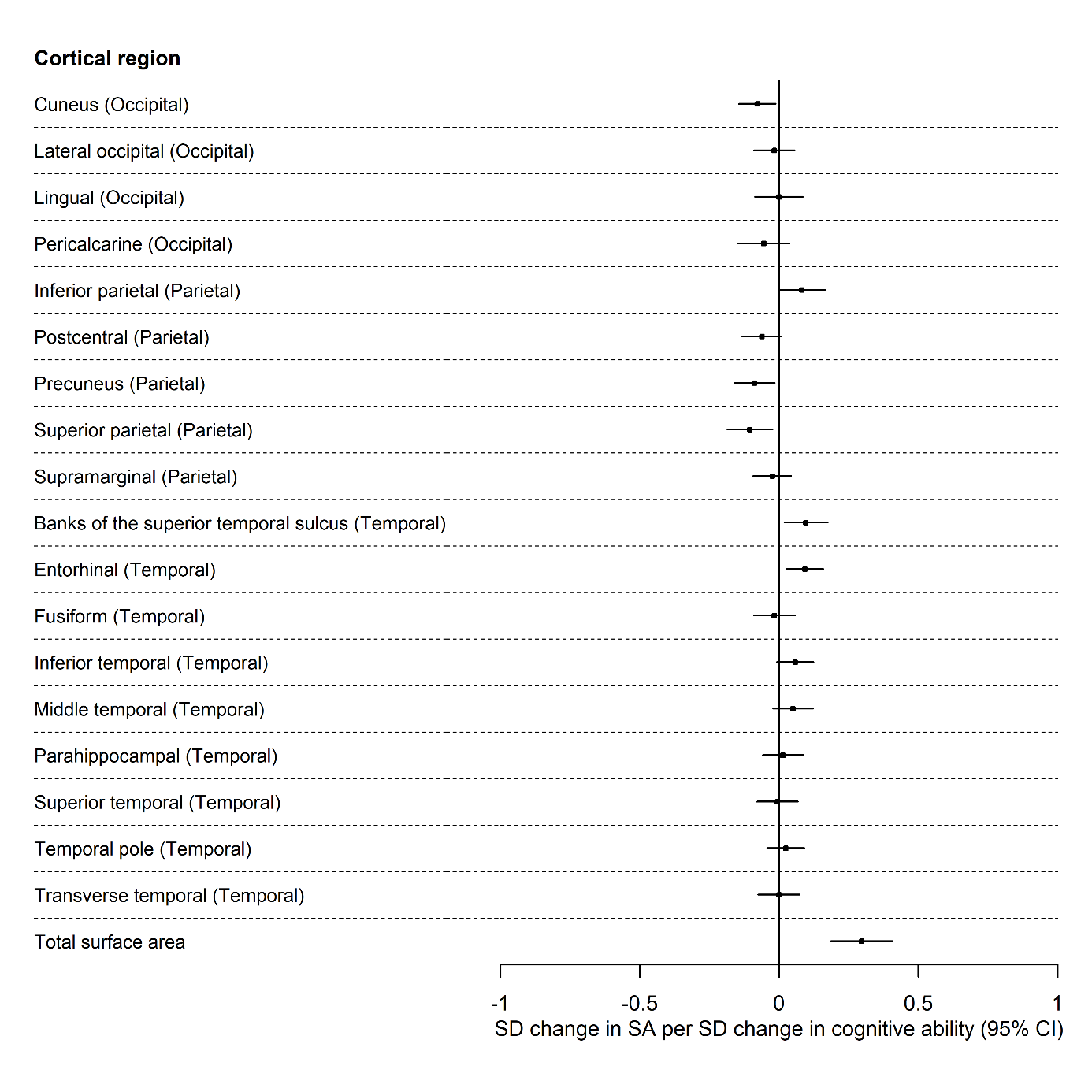

**Figure 2.2** The causal effects of genetically predicted cognitive ability on the surface area of the occipital, parietal, and temporal cortices and total surface area from the ENIGMA consortium.

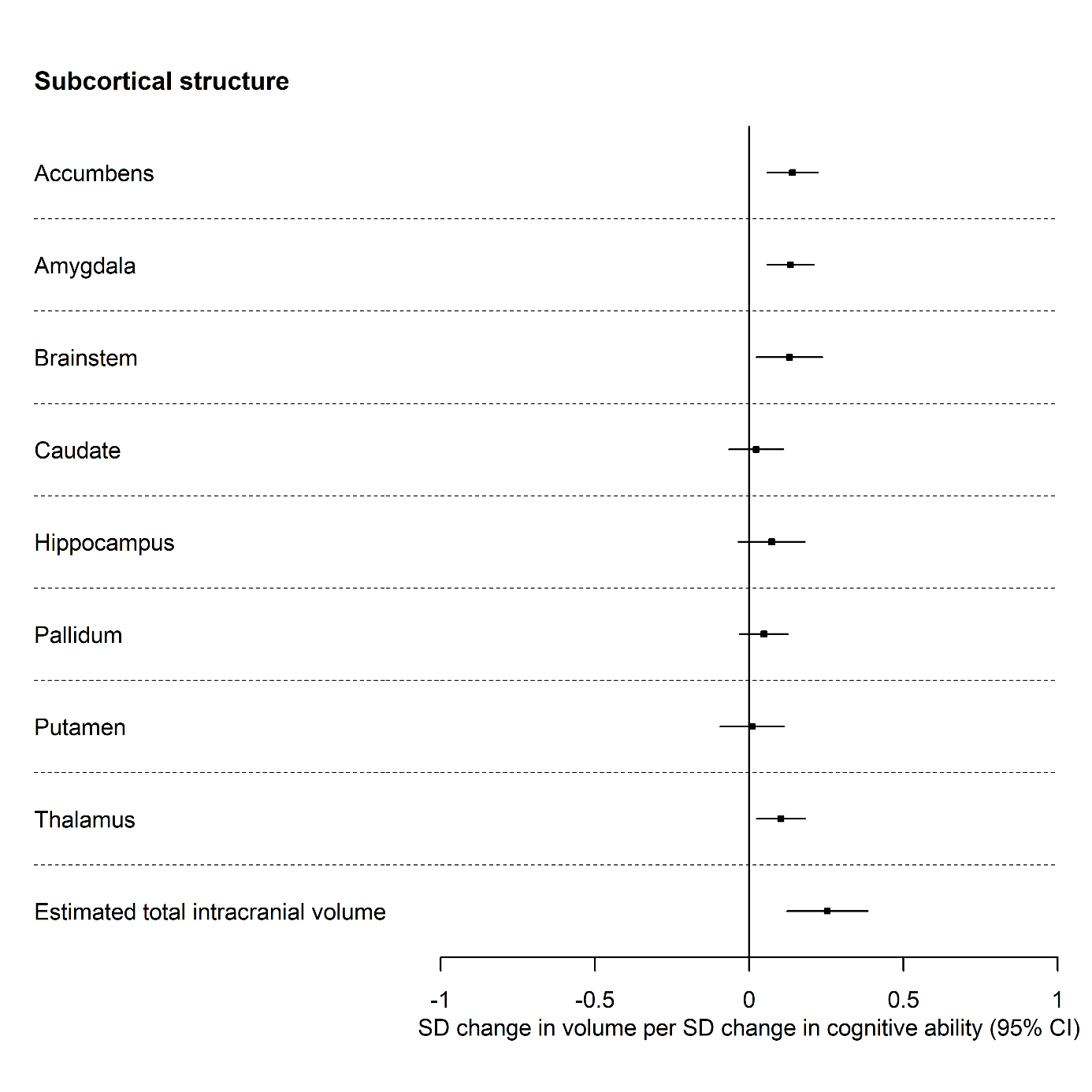

Figure 3 The causal effects of genetically predicted cognitive ability on the volume of subcortical structures from the ENIGMA consortium.

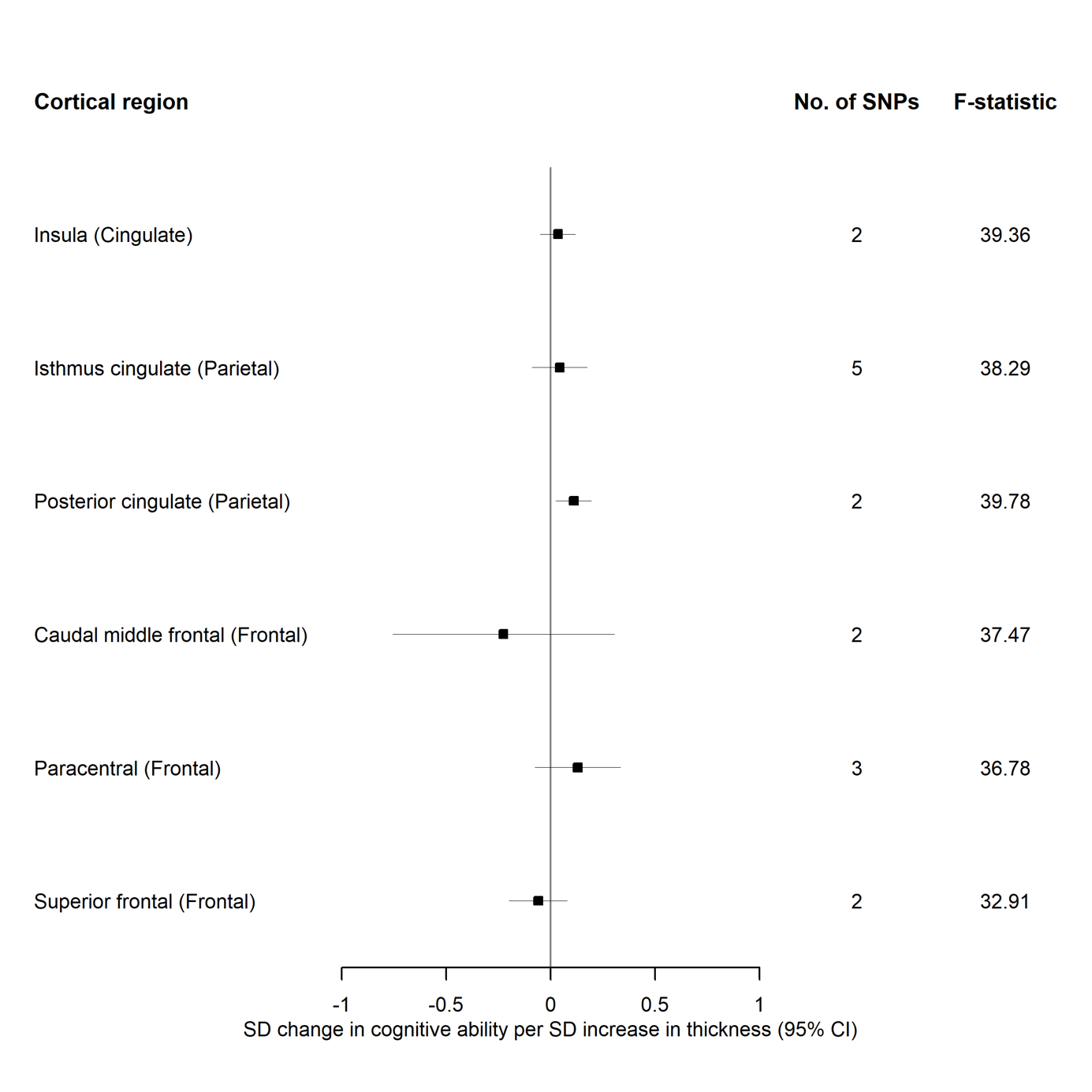

Figure 4.1 The causal effects of genetically predicted thickness of the cingulate, and frontal cortices from the ENIGMA consortium on cognitive ability.

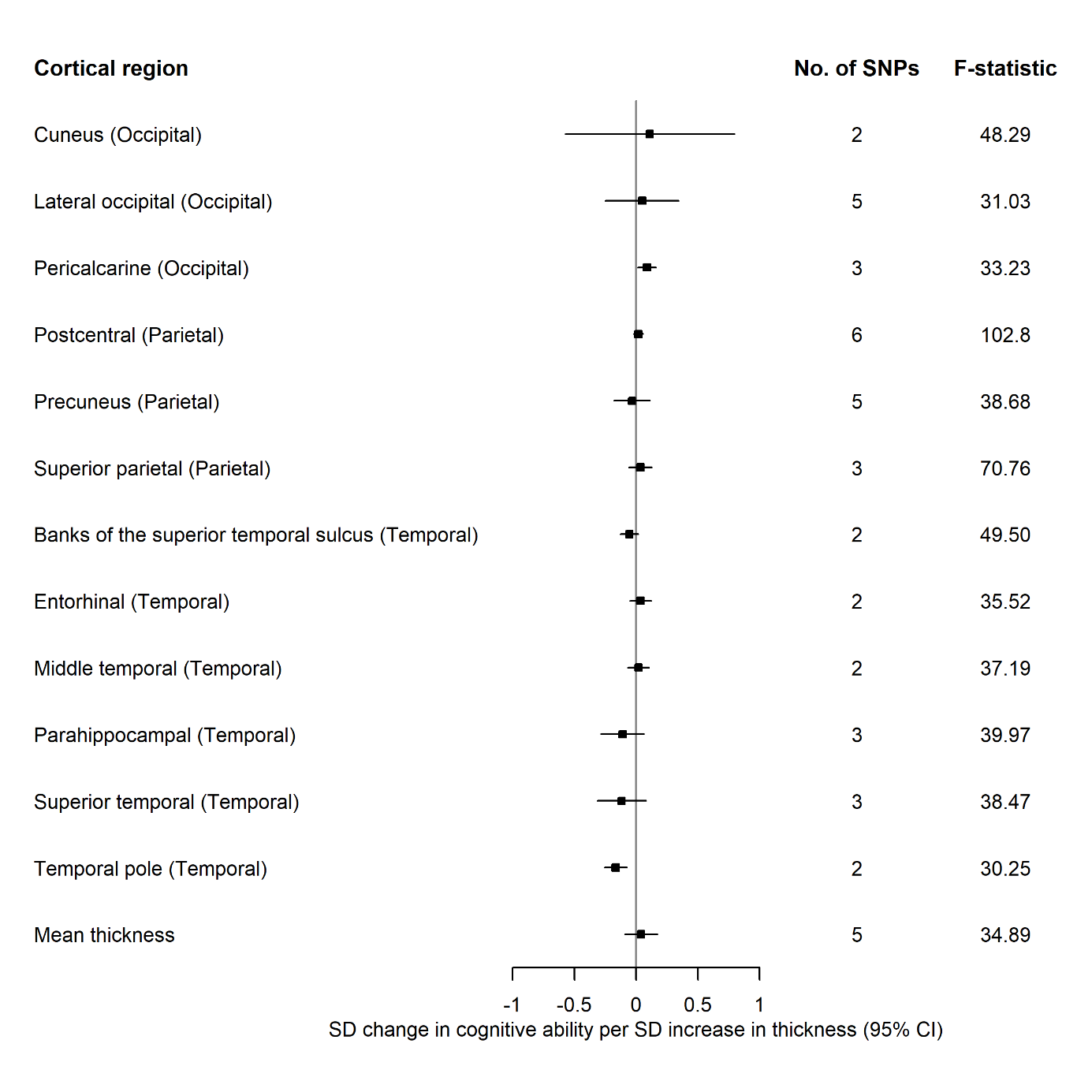

Figure 4.2 The causal effects of genetically predicted thickness of the occipital, parietal, and temporal cortices and mean thickness from the ENIGMA consortium on cognitive ability

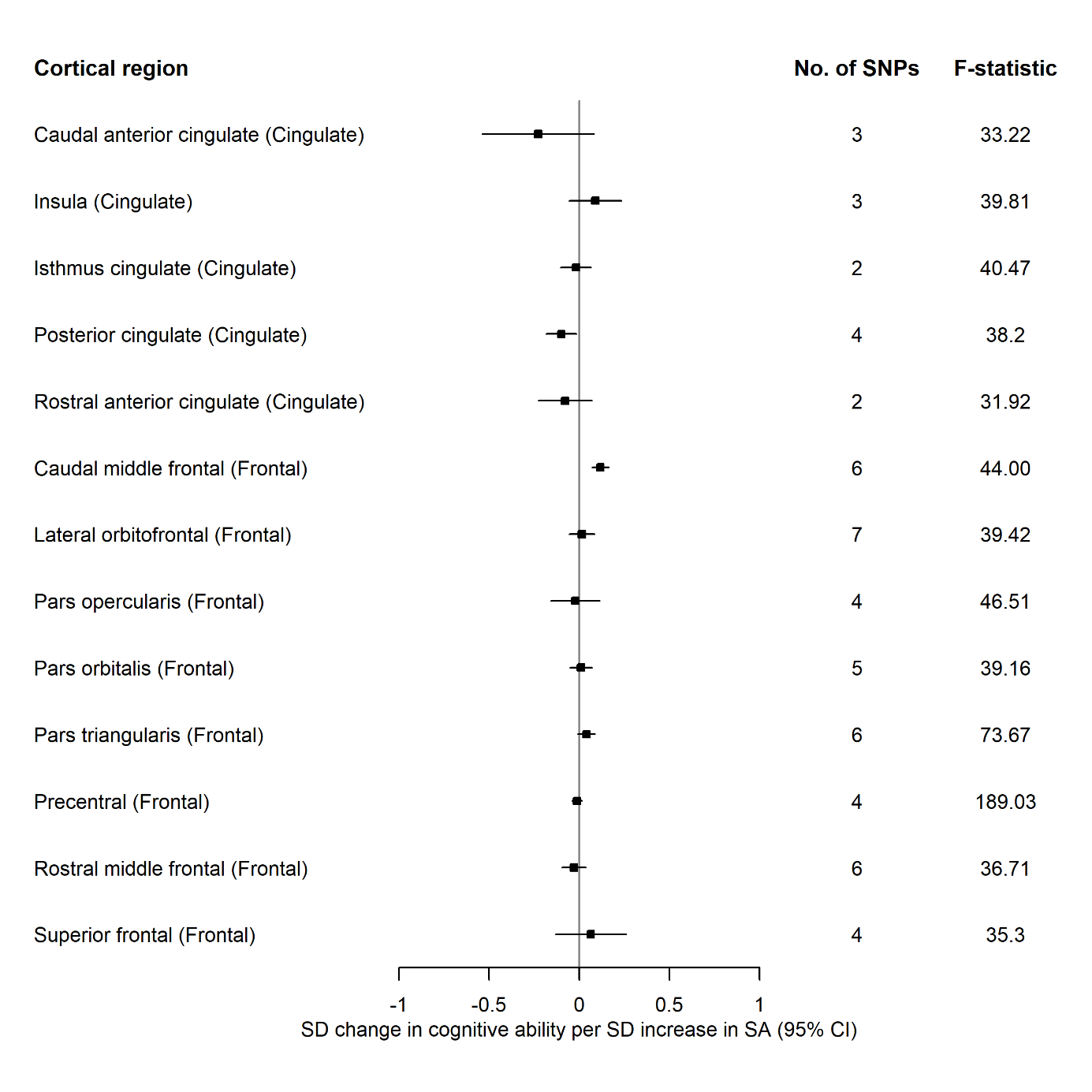
**Figure** **5.1** The causal effects of genetically predicted surface area of the cingulate, and frontal cortices from the ENIGMA consortium on cognitive ability. Abbreviations: SA, surface area**.**
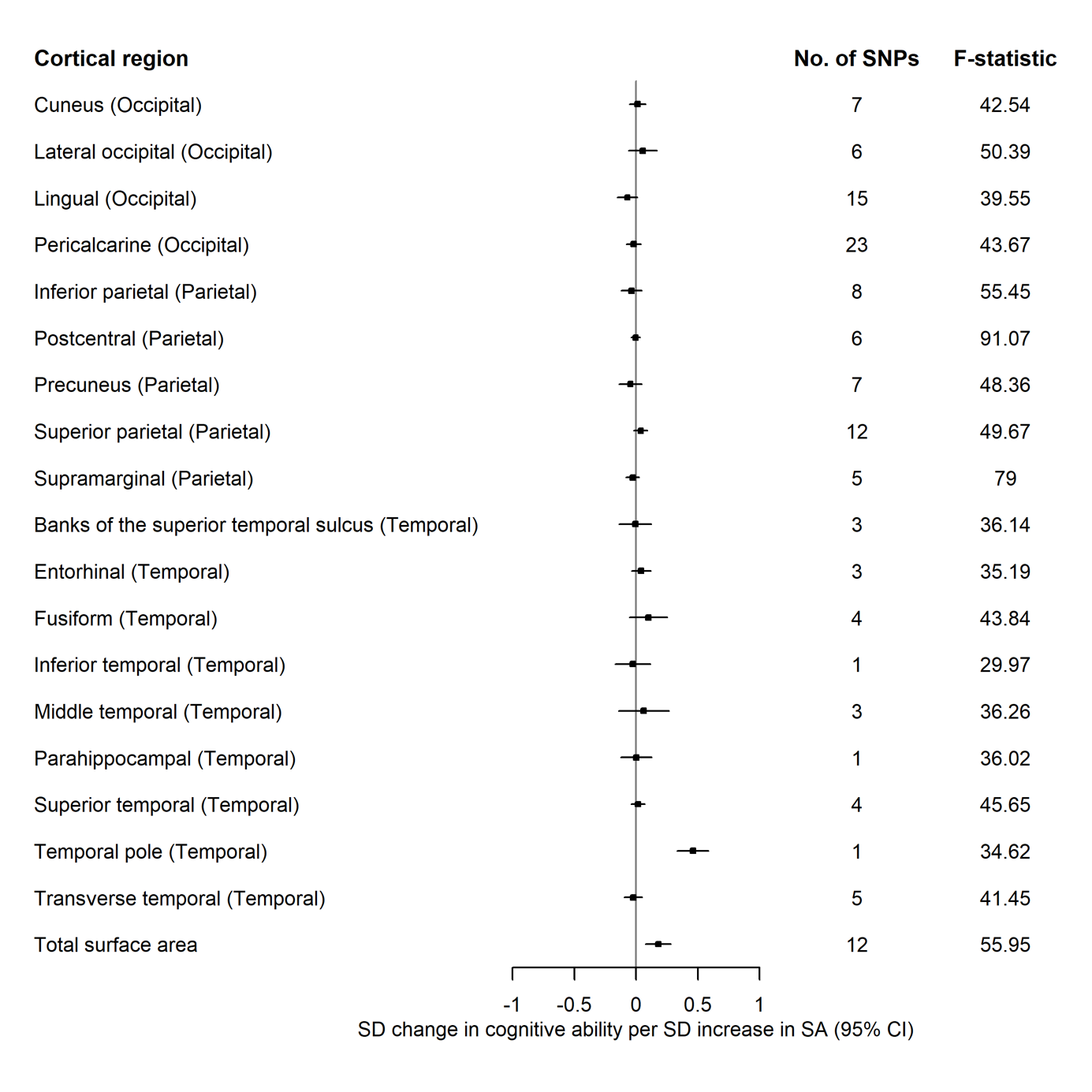

Figure 5.2 The causal effects of genetically predicted surface area of the occipital, parietal, and temporal cortices and total surface area from the ENIGMA consortium on cognitive ability. Abbreviations: SA, surface area

| **Table 1. Directionality test for the Mendelian randomization of cognitive ability on total surface area and estimated total intracranial volume in UK Biobank (age-stratified)** | | | | | | |
| --- | --- | --- | --- | --- | --- | --- |
| Tertile | Exposure | R^2^ for cognitive ability | R^2^ for exposure | Steiger p-value | SNPs removed* | IVW ^†(^95% CI) |
| 1 | Total surface area | 2.1% | 0.71% | 2.4x10^-9^ | 35 | 0.07 (-0.04, 0.18) |
| 2 |  | 1.9% | 0.58% | 2.08x10^-9^ | 51 | -0.03 (-0.14, -0.09) |
| 3 |  | 1.9% | 0.51% | 1.29x10^-10^ | 48 | 0.10 (-0.02, 0.21) |
| 1 | Estimated total intracranial volume | 2.0% | 0.61% | 2.66X10^-8^ | 41 | 0.06 (-0.06, 0.17) |
| 2 |  | 1.9% | 0.58% | 2.08x10^-9^ | 51 | -0.03 (-0.14, 0.09) |
| 3 |  | 1.8% | 0.41% | 9.92x10^-12^ | 54 | 0.08 (-0.04, 0.19) |

*SNPs which explain more variance in the outcome than the exposure. †IVW after removing SNPs with the false causal direction
